# Motherly Plus: Study Protocol of a Randomized Controlled Trial Evaluating a Smartphone-Assisted Single-Session Psychotherapy Intervention for Postpartum Depression

**DOI:** 10.64898/2026.11.18.26368427

**Authors:** Pedro F Zuccolo, Henrique Melo, Leonardo Seda, Tatiane Borja, Alicia Matijasevich, Arthur Caye, Euripedes Constantino Miguel, Renatha El Rafihi-Ferreira, Daniel Fatori

## Abstract

**Introduction:** Postpartum depression (PPD) is a public health problem, affecting approximately 17% of women worldwide. PPD is associated with significant costs to the healthcare system and impairments for the child. Although evidence shows that psychotherapy and pharmacotherapy are effective in reducing PPD symptoms, access to these treatments is limited, especially in low- and middle-income countries where there is a shortage of professionals and unequal distribution of mental health services. In this context, telehealth, smartphone apps, and brief psychotherapies emerge as viable, low-cost, and potentially scalable alternatives. However, gaps remain regarding the efficacy of single-session psychotherapies for PPD, especially when combined with digital support.

**Objectives:** The primary objective is to test the efficacy of an intervention package combining single-session psychotherapy with subsequent self-guided use of a smartphone app for the treatment of PPD. Secondary objectives are to investigate moderators, mediators, and specific intervention processes.

**Method:** A randomized clinical trial in which 104 women aged 18-40 years, up to 12 months postpartum, with a diagnosis of PPD will be randomized to the intervention (single-session psychotherapy supported by the Motherly app) or a waitlist. Participants in the intervention arm will retain access to the app throughout the study; waitlist participants will receive the intervention after post-treatment assessment. Assessments will occur at T1 (week 1, baseline), T2 (week 2, mid-treatment), T3 (week 4, post-treatment), and T4 (week 8, follow-up). The primary outcome will be depression assessed by the QIDS-C16 at T3. Secondary outcomes include anxiety, sleep, parental stress, and self-reported depression. Data will be analyzed using generalized estimating equations (GEE) models considering an intention-to-treat (ITT) perspective. Mediation models will be used to analyze mediators, while interaction models will be used to explore potential moderators. Regression models will be used to analyze the association between specific processes (therapeutic alliance and app engagement) and clinical outcomes.

**Perspectives:** This study has potential public health relevance and innovation by testing an intervention package that combines single-session psychotherapy and technology, with high potential for scalability.

**Trial registration:** clinicaltrials.gov NCT07925762

## Background

Postpartum depression (PPD) affects approximately 17% women worldwide (Wang et al., 2021), and its burden appears to have grown during the COVID-19 pandemic, particularly in low-resource settings (Chmielewska et al., 06/2021). PPD carries significant risks of chronicity (Jacques et al., 2020; Matijasevich et al., 2015), suicidality (Lindahl, Pearson, & Colpe, 2005), and adverse effects on child development (Rogers et al., 2020). PPD adds to the massive economic burden of mental health, estimated to cost £8.1 billion annually in the UK with 72% tied to child health and development issues, and an estimated R$26 billion in Brazil when accounting for hospital costs, lost productivity, reduced quality of life, and suicide-related expenses (Bauer, Knapp, Matijasevich, Osório, & de Paula, 2022; Bauer, Parsonage, Knapp, Iemmi, & Adelaja, 2014).

Current PPD treatment relies on pharmacotherapy and psychotherapy. Selective serotonin reuptake inhibitors (SSRIs) is the most frequent pharmacological choice, despite limited evidence and post-acute symptom remission (Brown et al., 2021). Novel PPD-specific medications, such as Zuranolone (Deligiannidis et al., 2023) and Brexanolone (Kanes et al., 2017), demonstrate efficacy, but lack regulatory approval in most countries, including Brazil. Ketamine and esketamine show promise for PPD prevention but are not yet recommended (Li, Zhou, Li, & Lin, 2024). Psychotherapy is efficacious, safe, and widely recommended (Cuijpers et al., 2023; Radoš et al., 2025). In low- and middle-income countries (LMICs), however, its implementation is hindered by a lack of culturally adapted protocols, shortages of trained professionals, and their uneven geographic distribution (Patel et al., 2018; Rathod et al., 2017; World Health Organization, 2021).

Aiming to address these barriers, our group developed Motherly, a smartphone app designed to offer psychotherapeutic strategies in an accessible format. Motherly Motherly combines tools for organizing maternal daily life with CBT-based mental health content, and behavioral activation (BA) is its main therapeutic component (Fatori et al., 2020). An earlier version was tested as an adjunct to a brief online CBT intervention for prenatal depression (Fatori, Zuccolo, Xavier, Matijasevich, & Polanczyk, 2023; Zuccolo, Xavier, Matijasevich, Polanczyk, & Fatori, 2021). Building on those findings, Motherly was redesigned and evaluated as a standalone intervention in a randomized controlled trial (RCT) with 264 women with PPD (Zuccolo et al., 2024). Compared with an active control app, Motherly did not significantly reduce depressive symptoms at post-treatment. It did produce small effects on sleep quality, parenting distress, BA, and clinician-rated improvement. Exploratory analyses suggested a dose–response relationship between app engagement and outcomes, but engagement was low, with a median of four BA activities registered during treatment (Zuccolo et al., 2024).

One hypothesis is that making therapeutic content available does not ensure that users can turn it into feasible actions in their daily lives. Women who used Motherly reported more activation than controls, but not greater access to environmental reward (Zuccolo et al., 2024). This suggests that the behavioral changes they made were not enough to produce meaningful reinforcement. Human support is known to increase engagement with digital interventions and their effects (Karyotaki et al., 2021; Mohr, Cuijpers, & Lehman, 2011). A single individual session before app use could help women select meaningful activities, anticipate barriers, adjust expectations to the realities of caring for an infant, and start using the app with guidance, while keeping the demand on professional resources low.

This model brings together three complementary modalities. Tele-mental health removes geographical barriers and has clinical efficacy comparable to face-to-face care (Scott et al., 2025; Singla, Silver, et al., 2025; Waugh, Mishkind, & Shore, 2018). Smartphone apps take advantage of high smartphone availability to deliver structured support after clinical contact ends (Goldberg, Lam, Simonsson, Torous, & Sun, 2022; Laricchia, 2023). Single-session psychotherapy concentrates care in one encounter, which reduces the impact of treatment attrition and the burden on professionals (Schleider et al., 2025). Single-session interventions have shown benefits for several mental health conditions, but the evidence carries a high risk of bias (Schleider et al., 2025). For PPD, it is limited to a few studies of group psychoeducational workshops delivered over several hours (Layton et al., 2025; Van Lieshout et al., 2021, 2023). To our knowledge, no trial has tested an individual single-session psychotherapy for PPD combined with digital support.

The Motherly Plus trial therefore asks what clinical benefit can be obtained from an intervention that concentrates scheduled psychotherapeutic contact in a single session and sustains practice through digital support. We will test whether a remote, BA-based single-session psychotherapy followed by four weeks of Motherly app use reduces depressive symptoms in women with PPD, compared with a waitlist. Furthermore, we will examine potential moderators of the intervention’s effect on the reduction of maternal depressive symptoms, such as age, income, education, marital status, social support, and the number of prenatal visits. We also intend to investigate potential mediation mechanisms, including changes in behavioral activation and access to reinforcers in the weeks following the intervention. Finally, we will analyze the association between treatment-specific processes, such as the therapeutic alliance and engagement with the app’s features, and the study outcomes.

## Methods

### Design

We will conduct a parallel-group randomized controlled trial (RCT) to test the efficacy of a smartphone app-supported single-session psychotherapy intervention. The study protocol was pre-registered on clinicaltrials.gov (NCT07925762). A total of 104 women (N=104) with postpartum depression (PPD), aged 18 to 40, will be randomized into two groups: (1) smartphone app-supported single-session psychotherapy, or (2) a waitlist control. Assessments will be conducted via the internet (remote interviews) and smartphone. The eligibility criteria are presented in Box 1, whereas Table 1 provides an overview of the RCT design.

**Box 1.**
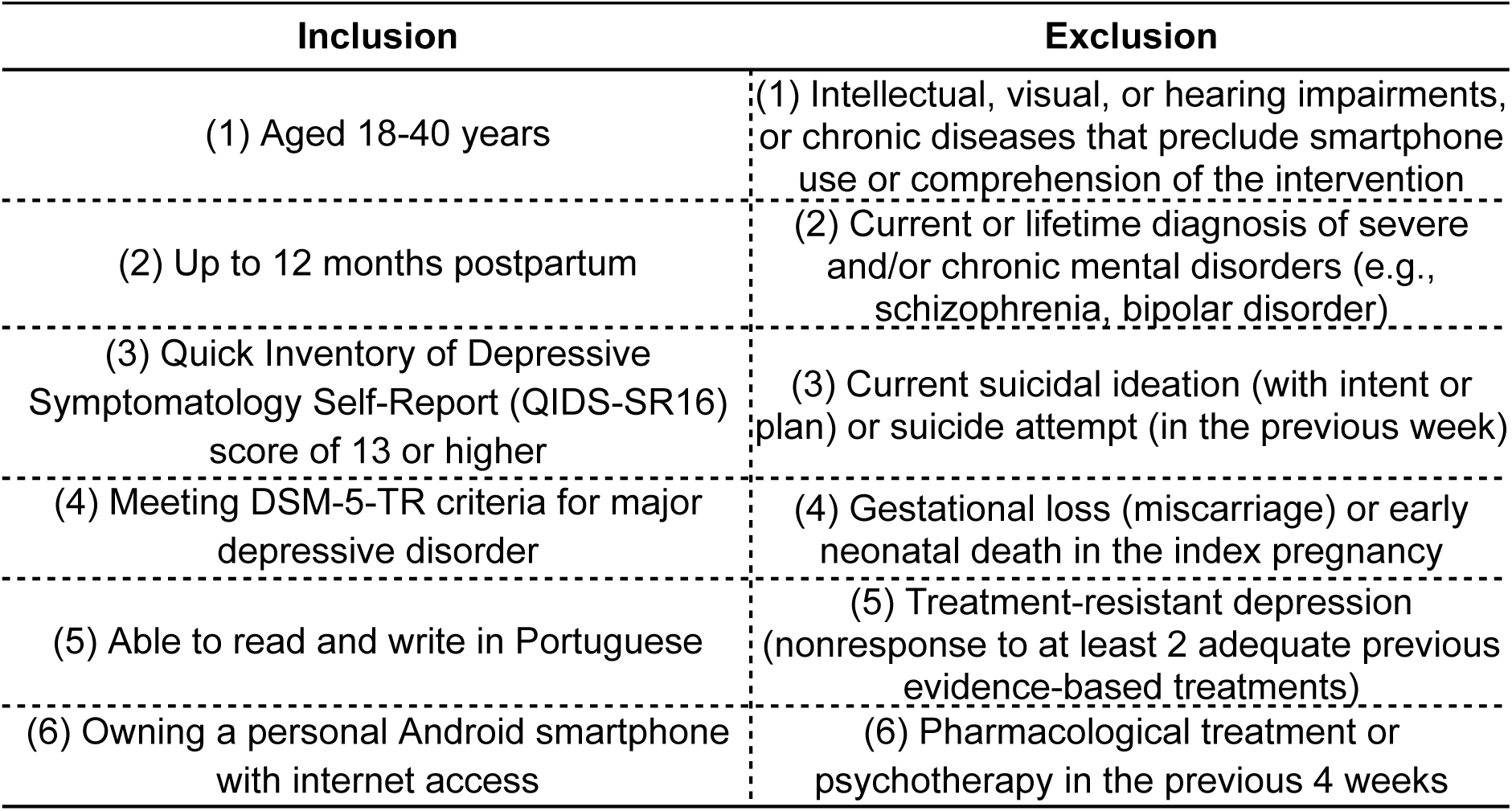
Eligibility criteria.

**Table 1.**
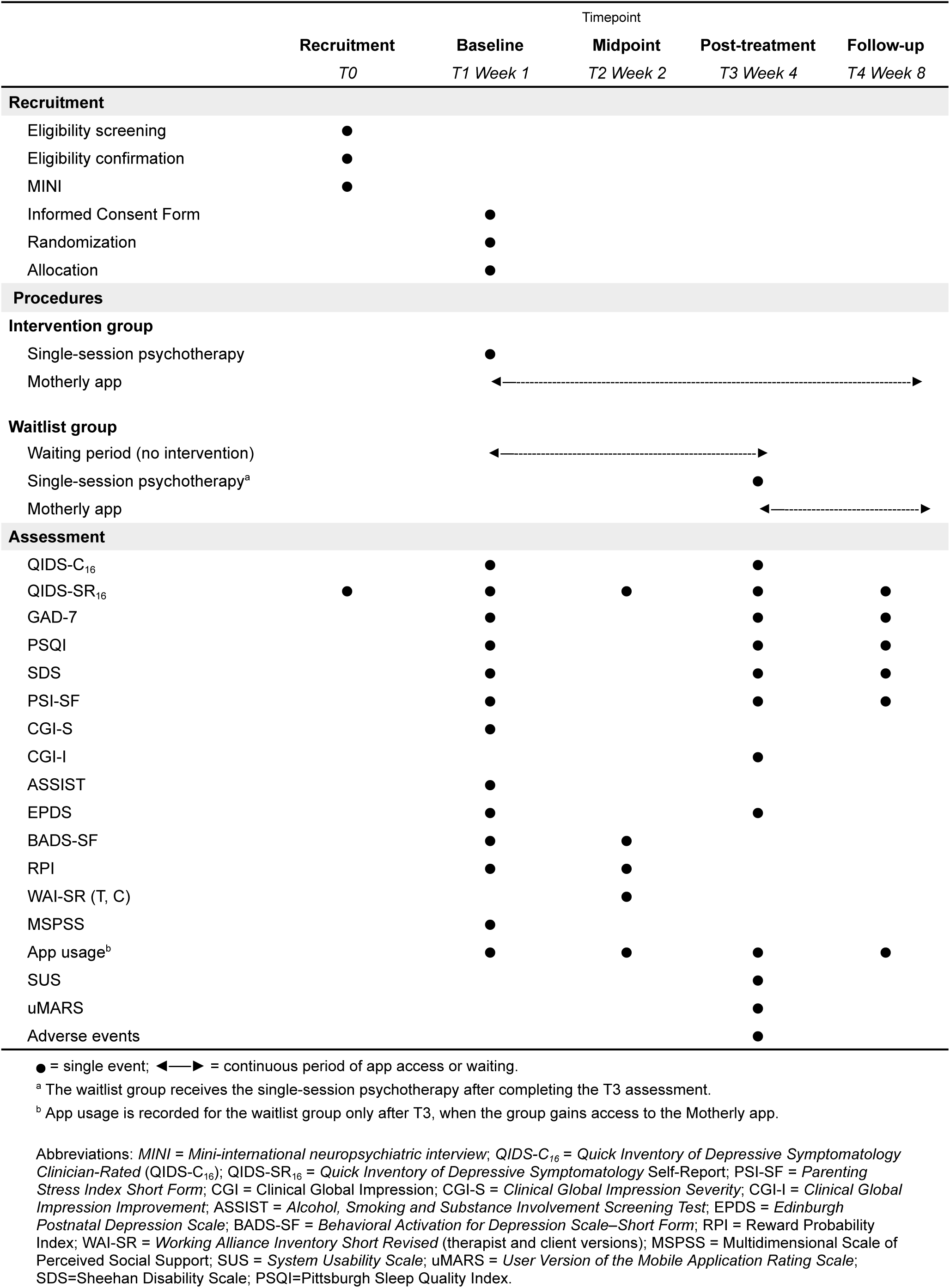
Overview of randomized controlled trial.

|  | Timepoint |  |  |  |  |
|---|---|---|---|---|---|
|  | **Recruitment** | **Baseline** | **Midpoint** | **Post-treatment** | **Follow-up** |
|  | *T0* | *T1 Week 1* | *T2 Week 2* | *T3 Week 4* | *T4 Week 8* |
| **Recruitment** |  |  |  |  |  |
| Eligibility screening | ● |  |  |  |  |
| Eligibility confirmation | ● |  |  |  |  |
| MINI | ● |  |  |  |  |
| Informed Consent Form |  | ● |  |  |  |
| Randomization |  | ● |  |  |  |
| Allocation |  | ● |  |  |  |
| **Procedures** |  |  |  |  |  |
| **Intervention group** |  |  |  |  |  |
| Single-session psychotherapy |  | ● |  |  |  |
| Motherly app |  | ◄──► |  |  |  |
| **Waitlist group** |  |  |  |  |  |
| Waiting period (no intervention) |  | ◄──► |  |  |  |
| Single-session psychotherapy<sup>a</sup> |  |  |  | ● |  |
| Motherly app |  |  |  | ◄──► |  |
| **Assessment** |  |  |  |  |  |
| QIDS-C<sub>16</sub> |  | ● |  | ● |  |
| QIDS-SR<sub>16</sub> | ● | ● | ● | ● | ● |
| GAD-7 |  | ● |  | ● | ● |
| PSQI |  | ● |  | ● | ● |
| SDS |  | ● |  | ● | ● |
| PSI-SF |  | ● |  | ● | ● |
| CGI-S |  | ● |  |  |  |
| CGI-I |  |  |  | ● |  |
| ASSIST |  | ● |  |  |  |
| EPDS |  | ● |  | ● |  |
| BADS-SF |  | ● | ● |  |  |
| RPI |  | ● | ● |  |  |
| WAI-SR (T, C) |  |  | ● |  |  |
| MSPSS |  | ● |  |  |  |
| App usage<sup>b</sup> |  | ● | ● | ● | ● |
| SUS |  |  |  | ● |  |
| uMARS |  |  |  | ● |  |
| Adverse events |  |  |  | ● |  |

● = single event; ◄──► = continuous period of app access or waiting.
<sup>a</sup> The waitlist group receives the single-session psychotherapy after completing the T3 assessment.
<sup>b</sup> App usage is recorded for the waitlist group only after T3, when the group gains access to the Motherly app.
Abbreviations: *MINI = Mini-international neuropsychiatric interview*; *QIDS-C<sub>16</sub> = Quick Inventory of Depressive Symptomatology Clinician-Rated* (QIDS-C<sub>16</sub>); QIDS-SR<sub>16</sub> = *Quick Inventory of Depressive Symptomatology* Self-Report; PSI-SF = *Parenting Stress Index Short Form*; CGI = Clinical Global Impression; CGI-S = *Clinical Global Impression Severity*; CGI-I = *Clinical Global Impression Improvement*; ASSIST = *Alcohol, Smoking and Substance Involvement Screening Test*; EPDS = *Edinburgh Postnatal Depression Scale*; BADS-SF = *Behavioral Activation for Depression Scale–Short Form*; RPI = Reward Probability Index; WAI-SR = *Working Alliance Inventory Short Revised* (therapist and client versions); MSPSS = Multidimensional Scale of Perceived Social Support; SUS = *System Usability Scale*; uMARS = *User Version of the Mobile Application Rating Scale*; SDS=Sheehan Disability Scale; PSQI=Pittsburgh Sleep Quality Index.

We plan to recruit participants in two stages. Stage 1 consists of completing an online electronic form with a checklist including inclusion criteria (1), (2), (3), (5), and (6), as well as exclusion criteria (1), (3), and (6). Stage 2 consists of: a) confirmation via telephone interview or video call for women who meet the inclusion criteria from Stage 1; and b) diagnostic assessment using the Mini-international Neuropsychiatric Interview (MINI) (Amorim, 2000; Sheehan et al., 1998) to confirm inclusion criterion (4) and exclusion criterion (2).

Women who have their inclusion criteria confirmed will be invited to a baseline assessment, which includes sociodemographic questions, a survey of risk factors, illnesses and disabilities, symptoms of depression and anxiety, sleep problems, parental stress, and the presence of concurrent treatments.

Throughout the study, both groups will answer questions regarding clinical symptoms of depression, anxiety, and sleep, among others (see Outcomes, Table 4). This information will be available in real time for researchers to assess whether participants require referral due to severe depressive symptoms or suicidal ideation. In such cases, the participant will be notified via message and telephone. Assessments will be conducted at baseline (T1, week 1), mid-treatment (T2, week 2), post-treatment (T3, week 4), and follow-up (T4, week 8).

### Intervention

The intervention consists of an online single-session psychotherapy protocol lasting approximately 90 minutes and based primarily on behavioral activation (BA), followed by self-guided use of a smartphone app developed to treat maternal depression and other mental health symptoms (Motherly app). Participants retain access to the app throughout the study; the primary outcome is assessed four weeks after the session (T3). The intervention was designed to promote in a scalable way two of the basic mechanisms thought to be responsible for the reduction in depressive symptoms through BA: increased contact with positive reinforcers and reduction of avoidance behaviors that limit access to reinforcing activities.

The single psychotherapy session focuses on the following components: psychoeducation about the behavioral model of depression, identification of sources of reinforcement and avoidance, collaborative identification and planning of value-aligned activities to interrupt avoidance and promote reinforcement, and training to use the app for implementing strategies in the following weeks.

Pre-session questionnaire: participants complete a structured questionnaire before the psychotherapy session, reflecting on their daily routine, sources of reinforcement, support networks, and activities they have discontinued since symptom onset. This component optimizes session time by prompting early engagement with treatment targets and allowing the therapist to tailor the intervention to the participant’s specific context.

Introduction and assessment: the session begins with rapport-building using active listening and psychoeducation about the single-session approach, followed by a focused assessment of the participant’s current daily activities, support networks, historical sources of reinforcement, prior experiences, and key life domains (e.g., child care, work, relationships). These foundational steps establish a collaborative relationship and gather information necessary for selecting meaningful behavioral activation targets.

Behavioral activation: the core intervention involves collaboratively identifying potentially reinforcing activities that are achievable in the participants’ current context, and scheduling them for the coming weeks. The therapist provides a clear, accessible explanation of how reducing activity and withdrawal maintain depression, and how increasing contact with reinforcing activities can interrupt this cycle. During this stage, the therapist identifies possible barriers to the implementation of the strategies and explores avoidance behaviors that decrease the probability of the participant actually engaging in reinforcing activities. Rather than targeting complete elimination of avoidance, the therapist helps the participant develop realistic, incremental strategies to gradually reduce these patterns. Therefore, the therapist focuses on small, manageable steps that are feasible within a single-session context. This emphasis on achievable, contextually-appropriate activities or reduction of avoidance behaviors reflects core single-session psychotherapy principles: leveraging existing resources, activating social support networks, recognizing the participant’s inherent capacity for change, and acknowledging that one session cannot resolve all difficulties but can be useful to produce meaningful changes. By focusing on realistic, feasible changes rather than idealized goals, the intervention builds on existing strengths and increases the likelihood of sustained engagement and success beyond the therapeutic encounter.

Additional CBT techniques: in addition to BA, our protocol will include optional components to be used according to the individual needs of the participant and the psychotherapist’s clinical judgment. Specifically, stress management focused on distress tolerance, emotion regulation techniques, cognitive restructuring, and sleep hygiene techniques.

Introduction to the Motherly app: the therapist guides the participant through using the Motherly app, which was developed to treat maternal depression and other mental symptoms using evidence-based components of CBT, as well as to function as a hub to organize the mother’s life during gestation or the postpartum period.

Motherly offers six mental health intervention modules, presented as interactive experiences organized into stages or phases, in which participants can learn specific strategies for managing mental health symptoms. The content is provided through short texts and graphic resources (e.g., illustrations, diagrams, etc.). Some stages also include input fields where participants can interact by entering their own experiences (e.g., behaviors, emotions, impressions, etc.) according to specific instructions aimed at practicing CBT techniques. These modules can be completed by participants at any time and at their own pace.

1. BA: This module offers psychoeducational content on how to implement BA techniques to increase engagement in reinforcing activities and solve problems that prevent access to reinforcing stimuli. App features support the implementation of BA, such as activity planning, mood and anxiety tracking, and progress monitoring.
2. Emotional Regulation: Based on the principles of Dialectical Behavior Therapy (DBT) (Leahy, Tirch, & Napolitano, 2011; Linehan, 2015), this module helps participants identify, label, and accept their emotions as normal, assisting in the development of effective strategies to manage negative emotions.
3. Stress Management: Also based on DBT principles (Leahy et al., 2011; Linehan, 2015), this module offers educational content on distress tolerance skills, including reality acceptance and breathing exercises that reduce autonomic arousal.
4. Cognitive Restructuring: This module provides interactive educational content aimed at helping participants identify and change dysfunctional thoughts and beliefs (Beck, 2021), leading to an adaptive perception of reality in accordance with CBT.
5. Sleep Hygiene and Relaxation: Utilizing evidence-based techniques developed to improve sleep quality (Murawski, Wade, Plotnikoff, Lubans, & Duncan, 2018; Perlis, Aloia, & Kuhn, 2010), this module offers psychoeducation on habits and practices that can enhance sleep quality, along with audio-guided psychoeducation on relaxation techniques to reduce somatic and cognitive arousal.
6. Mindfulness: Based on CBT-oriented mindfulness (Crane et al., 2017) and a program previously tested in Brazil (Pasqualucci, Seifert, Filho, & Dos Santos, 2022), this module offers audio lessons on the importance of mindfulness and audio-guided meditation sessions on how to meditate properly.

In addition to the mental health modules, the Motherly app includes features to help participants organize various aspects of daily life, such as scheduling health-related events (e.g., prenatal appointments or exams) and recording relevant information (e.g., weight, gestational age, depression symptoms). It also offers a library of educational content on a wide range of topics related to physical and mental health and child development. Table 2 summarizes the Motherly app features.

**Table 2.** Motherly app features.

| Module / feature | Focus | Objective / mechanism |
|---|---|---|
| **Mental health modules** |  |  |
| Behavioral Activation | Behavioral change and mood monitoring | Scheduling and structuring pleasant and productive activities and solving problems that limit access to reinforcement; supported by activity planning, mood tracking, and progress monitoring. |
| Emotional Regulation | Effective modulation of emotions | Identifying, labelling, normalizing, and accepting emotions (positive and negative); developing adaptive strategies to manage negative emotions. |
| Stress Management | Tolerance of painful emotions | Teaching distress tolerance skills, including reality acceptance and breathing exercises to reduce autonomic arousal. |
| Cognitive Restructuring | Identification and modification of thoughts | Identifying and disputing automatic thoughts and cognitive distortions, central mechanisms in CBT for depression and anxiety. |
| Sleep Hygiene and Relaxation | Sleep quality and duration | Psychoeducation on habits and practices that improve sleep; audio-guided relaxation techniques to reduce somatic and cognitive arousal. |
| Mindfulness | Present-moment awareness | Audio lessons on mindfulness and audio-guided meditation sessions, based on CBT-oriented mindfulness. |
| **Self management and information tools** |  |  |
| Pregnancy | Pregnancy monitoring and organization | Scheduling and organizing health-related events, such as prenatal appointments and exams, and tracking gestational age. |
| Assessments | Self-monitoring | Recording health information (e.g., weight) and mental health symptoms. |
| Library | Psychoeducation | Educational content on physical health, mental health, and child development, covering both pregnancy and the postpartum period. |

Abbreviations: CBT = cognitive behavioral therapy.

Through the Motherly app, specifically via the BA module described above, participants will be guided on how to schedule the agreed-upon behavioral activation activities discussed during the single-session psychotherapy. Participants can use the app’s scheduling system to plan activities, as well as monitor their progress regarding activity completion, symptom tracking, and overall progress monitoring. At any time, participants can also review psychoeducational content on BA to clarify doubts or improve their understanding of the concepts. Psychotherapists will also introduce additional psychoeducational modules (e.g., sleep hygiene, emotional regulation, mindfulness, postpartum-specific content), emphasizing the ones that are most appropriate for each participant according to their needs.

Providing technological support during the session facilitates engagement and ensures the participant can autonomously use the app for continued support over the subsequent weeks.

Session closing and commitment: the session concludes with a summary of key takeaways, review of the behavioral activation plan, and explicit discussion of how to maintain engagement despite obstacles. This consolidation strengthens treatment retention and reinforces the participant’s commitment to implement the strategies.

Self-guided app use: participants will use the Motherly app independently, scheduling and tracking behavioral activation activities, monitoring symptom progression, and accessing psychoeducational content as needed. Access to the app will remain available throughout the study, including the period between post-treatment (T3) and follow-up (T4). No additional therapist contact is planned.

The intervention will be delivered by psychotherapists with a degree in psychology. The selected psychotherapists will undergo 10 hours of training involving both theoretical and practical classes. During the study, the sessions will be supervised by senior researchers (P.F.Z., D.F.).

Table 3 provides an overview of each intervention component. For detailed information on intervention procedures, see the intervention manual (Supplementary Materials).

**Table 3.**
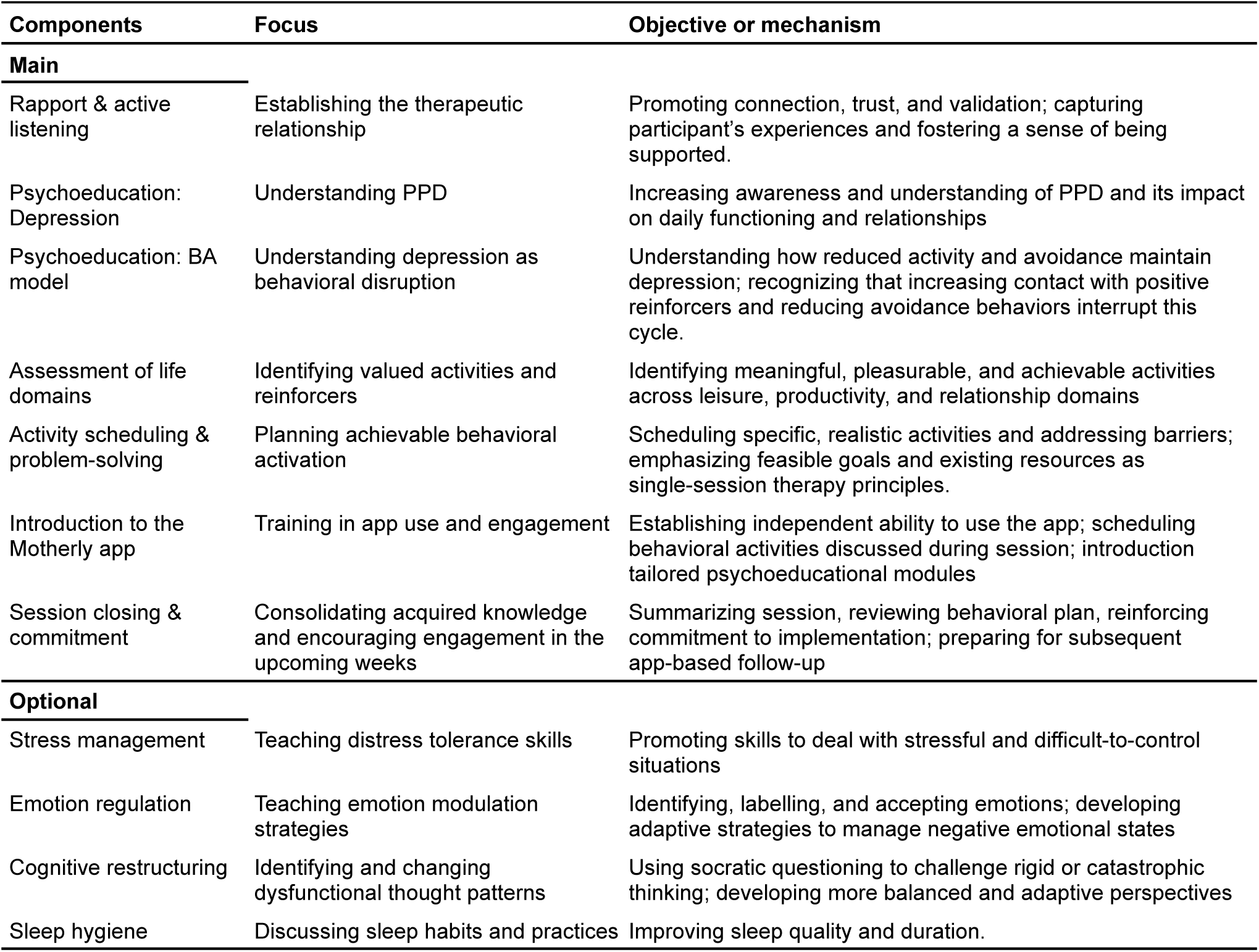
Intervention components and their objectives or putative mechanisms.

| Components | Focus | Objective or mechanism |
|---|---|---|
| **Main** |  |  |
| Rapport & active listening | Establishing the therapeutic relationship | Promoting connection, trust, and validation; capturing participant’s experiences and fostering a sense of being supported. |
| Psychoeducation: Depression | Understanding PPD | Increasing awareness and understanding of PPD and its impact on daily functioning and relationships |
| Psychoeducation: BA model | Understanding depression as behavioral disruption | Understanding how reduced activity and avoidance maintain depression; recognizing that increasing contact with positive reinforcers and reducing avoidance behaviors interrupt this cycle. |
| Assessment of life domains | Identifying valued activities and reinforcers | Identifying meaningful, pleasurable, and achievable activities across leisure, productivity, and relationship domains |
| Activity scheduling & problem-solving | Planning achievable behavioral activation | Scheduling specific, realistic activities and addressing barriers; emphasizing feasible goals and existing resources as single-session therapy principles. |
| Introduction to the Motherly app | Training in app use and engagement | Establishing independent ability to use the app; scheduling behavioral activities discussed during session; introduction tailored psychoeducational modules |
| Session closing & commitment | Consolidating acquired knowledge and encouraging engagement in the upcoming weeks | Summarizing session, reviewing behavioral plan, reinforcing commitment to implementation; preparing for subsequent app-based follow-up |
| **Optional** |  |  |
| Stress management | Teaching distress tolerance skills | Promoting skills to deal with stressful and difficult-to-control situations |
| Emotion regulation | Teaching emotion modulation strategies | Identifying, labelling, and accepting emotions; developing adaptive strategies to manage negative emotional states |
| Cognitive restructuring | Identifying and changing dysfunctional thought patterns | Using socratic questioning to challenge rigid or catastrophic thinking; developing more balanced and adaptive perspectives |
| Sleep hygiene | Discussing sleep habits and practices | Improving sleep quality and duration. |

### Control

Participants in the control group will be placed on a waitlist to receive the app-supported single-session psychotherapy intervention after completing the post-treatment assessment (T3, week 4). From then on, they will have access to the Motherly app under the same conditions as the intervention group. Below is an explanation of the rationale for using a waitlist as a comparison group.

The selection of a control condition in an RCT depends on factors such as the development phase of the intervention and the purpose of the study (Mohr et al., 2009). In early-stage studies (phase 1 or 2) of an intervention, a waitlist can be an appropriate choice to obtain a first impression of the intervention’s efficacy (Gold et al., 2017a). Although single-session psychotherapy has been discussed since the 1990s (Talmon, 1990), few RCTs have been conducted to test this intervention in adults with depression (Kim, Ryu, & Chibanda, 2023a). Therefore, the present RCT is considered a phase 2 study (evaluation in a larger group of patients to test efficacy and further investigate safety) (Gold et al., 2017a). Furthermore, the RCTs that have tested single-session psychotherapy already utilized control groups such as waitlists and no-treatment; however, all of these presented a high risk of bias according to the risk-of-bias tool (RoB2) (Kim et al., 2023a). Thus, this RCT contributes by producing evidence with a low risk of bias, robustly consolidating the effect of this intervention modality when compared to a waitlist control group. Finally, it is worth noting that a waitlist is not indicated for high-risk participants, such as those with a high probability of suicide attempt; therefore, the presence of current suicidal ideation, as well as refractory depression, are part of the exclusion criteria for the present study.

### Outcomes

Table 4 presents the primary and secondary outcomes of the study. Each outcome is described below.

**Table 4.** Primary and secondary outcomes.

| **Outcome** | **Instrument** | **Timepoint** |
|---|---|---|
| **Primary outcome** |  |  |
| Depression symptoms | QIDS-C<sub>16</sub> | T3 |
| **Secondary outcomes** |  |  |
| Depression symptoms | QIDS-SR<sub>16</sub>, EPDS | T3, T4 |
| Anxiety symptoms | GAD-7 | T3, T4 |
| Sleep problems | PSQI | T3, T4 |
| Parenting stress | PSI-SF | T3, T4 |
| Functioning | SDS | T3, T4 |
| Depression severity | CGI-I | T3 |

Abbreviations: T3 = 4 weeks, post-treatment; T4 = 8 weeks, follow-up. *QIDS-C<sub>16</sub> = Quick Inventory of Depressive Symptomatology Clinician-Rated* (QIDS-C<sub>16</sub>); QIDS-SR<sub>16</sub> = *Quick Inventory of Depressive Symptomatology* Self-Report; EPDS=Edinburgh Postnatal Depression Scale; GAD-7=Generalized Anxiety Disorder-7; PSQI=Pittsburgh Sleep Quality Index; PSI-SF = *Parenting Stress Index Short Form*; SDS=Sheehan Disability Scale; CGI-I=Clinical Global Impression Improvement.

#### Primary outcome

The primary outcome of the study will be depressive symptoms, operationalized as the total score of the Quick Inventory of Depressive Symptomatology Clinician-Rated (QIDS-C16) at post-treatment (T3, week 4).

#### Secondary outcomes

Depressive symptoms, defined as the total score of the Quick Inventory of Depressive Symptomatology Self-Report (QIDS-SR16) at post-treatment (T3, week 4) and follow-up (T4, week 8).

Depressive symptoms, defined as the total score of the Edinburgh Postnatal Depression Scale (EPDS) at post-treatment (T3, week 4) and follow-up (T4, week 8).

Anxiety symptoms defined as the total score of the GAD-7 at post-treatment (T3, week 4) and follow-up (T4, week 8).

Sleep problem symptoms, defined as the total score of the PSQI at post-treatment (T3, week 4) and follow-up (T4, week 8).

Parental stress symptoms, defined as the scores for parental distress, parent-child dysfunctional interaction, and total stress from the Parenting Stress Index Short Form (PSI-SF) at post-treatment (T3, week 4) and follow-up (T4, week 8).

Functional impairment, defined as the total score of the Sheehan Disability Scale (SDS) at post-treatment (T3, week 4) and follow-up (T4, week 8).

Change in depression severity and functional impairment relative to baseline (T1), defined as the score on the Clinical Global Impression Improvement (CGI-I) scale at post-treatment (T3, week 4).

Because waitlist participants receive the intervention after T3, between-group comparisons at T4 do not constitute a randomized comparison against an untreated control. T4 data will be used to examine maintenance of effects in the intervention group and within-group change in the waitlist group following delayed treatment.

### Moderators

A moderator is a variable that explains differential treatment outcomes, identifying which participant characteristics or under which circumstances an intervention yields better results (Hayes, 2022; Kraemer, Wilson, Fairburn, & Agras, 2002). By definition, a moderator is a variable measured at the study’s baseline. Identifying moderators provides support for personalized clinical treatment, allowing interventions to be recommended for individuals with specific profiles.

Because the sample size was computed based on the primary outcome (n = 104), the trial is not powered to detect treatment-by-moderator interactions, and all moderation analyses will therefore be considered exploratory and hypothesis-generating. The following variables will be explored as potential moderators in relation to the previously described outcomes: maternal age, maternal skin color/ethnicity, maternal education level, family income, number of previous children, relationship with the father, social support according to the Multidimensional Scale of Perceived Social Support (MSPSS), baby’s age, average time spent with the child, number of pediatric and prenatal consultations, baseline depression symptoms according to the QIDS-SR16 and QIDS-C16, baseline severity according to the CGI-S, and functionality according to the SDS.

### Mediators

A mediator is a variable that may help explain how an intervention influences an outcome, that is, a potential pathway through which the intervention operates (Kraemer, Kiernan, Essex, & Kupfer, 2008; Kraemer et al., 2002). By definition, a mediator is measured during treatment, and changes in the mediator should precede changes in the outcome (Kazdin, 2007). Understanding the mediators of an intervention allows for the advancement of knowledge regarding the intervention, not only validating it but also allowing for its optimization by focusing on the mechanisms that provide the greatest effect.

Evidence from RCTs suggests that BA-based interventions for maternal depression operate through changes in specific behavioral and interpersonal processes. For example, a secondary analysis of two RCTs found that increases in BA—defined as self-reported engagement in varied activities and the associated sense of pleasure or competence (mastery)—mediated the reduction of depressive symptoms (Singla et al., 2021). Furthermore, increases in perceived social support also acted as mediators of the intervention effect (Singla et al., 2021).

Similar results were observed in the Happy Mother, Healthy Baby (HMHB) trial, a structured psychosocial intervention based on CBT principles, including BA (A. Malik et al., 2024). In that study, social support and the reduction of gestational stressors (“pregnancy hassles”) mediated the effects on anxiety and depression in the postpartum period (A. Malik et al., 2024). These findings align with an RCT conducted in the USA by (Dimidjian et al., 01/2017), which demonstrated that early changes in BA and environmental reward mediated the improvement of depressive symptoms among pregnant women treated with BA. A recent systematic review reinforces the consistency of these mechanisms by showing that increased BA, reduced avoidance, strengthened social support, gains in emotional regulation, and greater psychological flexibility are associated with the therapeutic effects of perinatal psychotherapies (Singla, Lawson, et al., 2025). Finally, evidence from qualitative studies involving interviews and consultations with young people with lived experience of depression and anxiety indicates that women and young people consider BA an acceptable, concrete, and meaningful component of interventions (K. Malik et al., 2021).

Although there are studies investigating mediators of BA-based interventions for maternal depression, none of them involve a single-session psychotherapy followed by digital support. Thus, the processes involved in the effect of this type of intervention will be investigated. Specifically, the following measures will be used to investigate intervention mediators:

BA behaviors and avoidance, as defined by the total score of a modified version of the Behavioral Activation for Depression Scale–Short Form (BADS-SF) (Abreu & Abreu, 2020; Manos, Kanter, & Luo, 2011). In its original form, this scale examines BA and avoidance over the last seven days. The items will be reformulated to capture events occurring over the previous two weeks, so that each assessment covers the whole interval since the preceding time point.

Access to response-contingent positive reinforcement in the environment in the previous Two weeks, as defined by the total score of the Reward Probability Index (RPI).

Both mediators will be assessed at two time points: baseline (T1) and at midpoint (T2). Depressive symptoms will be assessed at baseline (T1), midpoint (T2), and post-treatment T3 to capture early change following the single session. This design addresses the requirement that changes in the mediator precede changes in the outcome (Kazdin, 2007). Because the modified BADS-SF covers the previous two weeks, the T2 assessment captures behavioral change across the interval between the single session and midpoint, which precedes the one-week recall window of the depressive symptom measure. Mediation models will use depressive symptoms at T3 as the outcome.

At the same time, symptom change in psychotherapy frequently occurs early in treatment, sometimes before the proposed mechanisms have been assessed (Kazdin, 2007). Depressive symptoms will therefore also be assessed at T2 to capture early change following the single session. If substantial improvement is already observed at T2, mediation models with T3 outcomes will be interpreted as testing whether behavioral processes account for subsequent change or maintenance of gains, rather than the initial treatment response.

The hypothesis is that the increase in BA and reduction in avoidance in the weeks following the single session will mediate the intervention’s effect on the reduction of depressive symptoms. Another hypothesis is that the increase in access to reinforcers, as measured by the RPI, will function as a mediator of the intervention. These hypotheses will be investigated using parallel mediation models (Hayes, 2022) adjusting for baseline (T1) levels of depressive symptoms and of the mediators. Because mediators are self-reported retrospectively and are not randomized, reports may be influenced by current mood, and mediation estimates will be interpreted with caution rather than as definitive evidence of causal mechanisms. Moreover, given the sample size, mediation analyses will be considered exploratory.

### Treatment-Specific Processes

Specific treatment processes are active components present only in the group receiving the intervention that may contribute to clinical change; however, they cannot be evaluated as formal mediators because they exist only in the study’s intervention group. They represent elements that help explain how the intervention exerts its effects among those who receive it, but they do not meet the statistical criteria necessary to characterize causal mediation. The following processes will be investigated.

Therapeutic alliance defined by the total scores of the Working Alliance Inventory Short Revised (WAI-SR), client and therapist versions, assessed at T2, two weeks after the single session. Because the intervention relies on a single therapeutic encounter, the alliance established in this session may be particularly relevant, as it could influence subsequent engagement in behavioral activation and with the digital support. We will therefore examine whether therapeutic alliance is associated with app engagement and with clinical outcomes.

Engagement and Motherly app usage measures, characterized by seven indicators: (1) completion of the BA tutorial; (2) number of BA activities scheduled in the app; (3) number of psychoeducational contents viewed; (4) count of screens accessed in the app; (5) count of modules accessed in the app; (6) app usage time; (7) number of audio tracks from the relaxation module listened to.

### Randomization and allocation

Randomization will be performed by an independent statistician using R. Participants will be automatically allocated to the intervention or control groups through the REDCap randomization module. Thus, there will be no interference or prior knowledge by the researchers, ensuring allocation concealment. Once participants are allocated, the study management will contact them to inform them of their allocation and schedule the necessary procedures.

### Recruitment

Participants will be identified through recruitment on social media and the project’s website. Women interested in participating in the study will be invited to complete a digital form containing questions related to the inclusion and exclusion criteria. Potential participants who meet the inclusion criteria will be contacted via telephone by a researcher from the study team to confirm the criteria, explain the study, and confirm their interest in participating. Women who wish to participate will be asked to digitally sign an informed consent form and will subsequently be randomized into either the intervention or the waitlist group.

### Assessments

A comprehensive clinical assessment of maternal physical and mental health, demographics, and psychological characteristics of the participants will be conducted throughout the study. The assessments will be carried out by clinicians specifically trained for this study. The training will consist of lectures with a total workload of twenty hours.

The assessors will be masked to the group to which the participants were randomized and allocated. All data will be entered electronically via Research Electronic Data Capture (REDCap) (Harris et al., 2009). The standardized instruments to be used in the study are listed below.

Quick Inventory of Depressive Symptomatology Clinician-Rated (QIDS-C16): The QIDS-C16 is a scale developed to investigate depression through an assessment conducted by a trained clinician (Reilly, MacGillivray, Reid, & Cameron, 2015; Rush et al., 2003a). The QIDS includes items covering all major depressive disorder criteria from the Diagnostic and Statistical Manual of Mental Health Disorders, Fifth Edition, Text Revision (DSM-5-TR) (American Psychiatric Association, 2023) and evaluates nine domains: sadness, concentration, self-criticism, suicidal ideation, general interest, energy/fatigue, sleep problems, appetite/weight, and psychomotor skills. All domains are covered by a single item, with the exception of sleep problems (four items), appetite/weight (four items), and psychomotor agitation/retardation (two items). The items with the highest score within these domains are added to the scores of the other items. The total score ranges from 0 to 27, with higher scores representing more symptoms. The interpretation of the scores is as follows: 0 to 5 for absence of depression, 6 to 10 for mild depression, 11 to 15 for moderate depression, 16 to 20 for severe depression, and 21 to 27 for very severe depression.

The rationale for selecting the QIDS-C16 as the primary outcome assessment instrument is based on the following reasons: (1) it provides a complete evaluation of the three primary psychopathological domains of major depressive disorder (Malhi & Mann, 2018): neurovegetative, emotional, and neurocognitive; (2) it offers a comprehensive assessment of all symptoms for major depressive disorder criteria according to the DSM-5-TR: sadness, concentration changes, negative self-criticism, suicidal ideation, interest (anhedonia), fatigue, sleep problems, appetite/weight, and psychomotor changes; and (3) the assessment of symptoms is conducted by a trained clinician who investigates in detail the presence and severity of each symptom. As a semi-structured interview with standardized introductory questions, each item is scored based on clinical judgment. This approach allows for a masked assessment; if the participant were the sole informant, bias could occur due to knowledge of the allocation in studies where the control is not a placebo or sham (Sterne et al., 2019a).

Quick Inventory of Depressive Symptomatology Self-Report (QIDS-SR_16_): The QIDS has a specific version for assessing depression through self-reporting. The QIDS-SR16 features the same domains, symptoms, and items as the QIDS-C_16_, with the only difference being the method of administration. The rationale for using the QIDS-SR_16_ is the need for a complementary subjective perspective of the participant’s depressive symptoms.

Edinburgh Postnatal Depression Scale (EPDS): The EPDS is a 10-item scale developed to assess pre- and postnatal depression. It is the most widely used scale globally in studies related to pregnancy and the postnatal period. It has been extensively used in epidemiological studies for screening purposes and in RCTs to measure symptom severity (Santos et al., 2007). The EPDS will be used for comparison with the QIDS, given that the EPDS is more common in the PPD literature.

It is worth noting that the EPDS will not be used as the primary outcome due to its limitations in capturing all symptomatic domains of PPD. The EPDS disproportionately emphasizes emotional symptoms, with six items focused on this domain, whereas only two address neurovegetative symptoms and two do not strictly constitute depressive symptoms. Furthermore, the scale completely lacks items addressing neurocognitive symptoms which are frequent in postpartum depression, such as diminished ability to think, concentrate, or indecisiveness (Radoš et al., 2024).

Generalized Anxiety Disorder scale (GAD-7): The GAD-7 is a brief 7-item scale used to assess anxiety disorders. Its brevity and simplicity allow for widespread use in primary care, primarily for screening anxiety problems in populations. It also enables a dimensional understanding of anxiety via a continuous score, thus serving as a measure of severity (Moreno et al., 2016; Plummer, Manea, Trepel, & McMillan, 2016).

Pittsburgh Sleep Quality Index (PSQI): Sleep quality over the past month is a fundamental indicator of physical and mental health. The PSQI scale assesses subjective quality and sleep efficiency, identifying difficulties through seven domains: subjective quality, latency, duration, habitual efficiency, sleep disturbances, use of medication, and daytime dysfunction (Bertolazi et al., 2011; Buysse, Reynolds, Monk, Berman, & Kupfer, 1989).

Parenting Stress Index Short Form (PSI-SF): Parental care in a child’s first months of life can generate stress related to various aspects, such as daily activities, support from relatives and partners, and interaction with the child, among others. The PSI-SF specifically assesses stress derived from parental care, allowing for an understanding of the presence or absence of stress levels across the following domains: parental distress, parent-child dysfunctional interaction, difficult child (behavioral problems), and total stress (the sum of the domains) (Pereira, Viera, Toso, Carvalho, & Bugs, 2016).

Sheehan Disability Scale (SDS): Depression can significantly compromise an individual’s functioning, leading to impairments in various areas of life. The SDS specifically assesses the disruption and functional disability caused by symptoms, allowing for the measurement of the level of impairment (from “not at all” to “extremely”) in the following domains: work/school, social life, and family life/home responsibilities (Sheehan et al., 1996). Previous studies have established a relationship between functionality according to the SDS and the severity of depression (Guico-Pabia, Fayyad, & Soares, 2012).

Clinical Global Impression (CGI) Severity (CGI-S) and Improvement (CGI-I): This set of scales constitutes a method for assessing the severity of an illness or mental disorder and the improvement in functioning in response to treatments (Guy, 1976). The CGI-S classifies individuals diagnosed with a mental disorder into seven categories (1, Normal, not at all ill; 2, Borderline mentally ill; 3, Mildly ill; 4, Moderately ill; 5, Markedly ill; 6, Severely ill; 7, Among the most extremely ill patients). Meanwhile, the CGI-I classifies the individual’s change into seven possibilities (1, Very much improved; 2, Much improved; 3, Minimally improved; 4, No change; 5, Minimally worse; 6, Much worse; 7, Very much worse) (Lima et al., 2007).

Alcohol, Smoking and Substance Involvement Screening Test (ASSIST): an instrument developed by the World Health Organization to assess the use, abuse, and dependence on alcohol, tobacco, and other substances (e.g., cocaine, anxiolytics, stimulants, etc.) (Henrique, De Micheli, Lacerda, Lacerda, & Formigoni, 2004). It allows for both dimensional and categorical assessment of substance use-related problems. A modified version of the ASSIST will be administered to verify current use of tobacco, alcohol, and other substances.

Behavioral Activation for Depression Scale–Short Form (BADS-SF): the BADS is a self-report scale designed to measure changes in BA and avoidance behaviors (Manos et al., 2011), consistent with the BA theory and treatments proposed by (Martell, Dimidjian, & Herman-Dunn, 2013). In its short form, it consists of 9 Likert-type items (ranging from 0 = completely disagree to 6 = completely agree), with scores ranging from 0 to 54. Higher scores indicate an increase in activation and a decrease in avoidance behaviors. In the original scale, the questions refer to the previous week; for this study, the items will be modified to refer to the previous two weeks.

Reward Probability Index (RPI): the RPI is a self-report scale that measures the probability of reinforcement and the presence of environmental suppressors, serving as a measure of environmental changes associated with Behavioral Activation (BA) treatment (Carvalho et al., 2011). It contains 20 Likert-type items, with scores ranging from 20 to 80. Higher scores indicate an increased probability of reward and fewer environmental suppressors inhibiting access to reinforcers.

Multidimensional Scale of Perceived Social Support 3–item version (MSPSS): the MSPSS will be used as a measure of maternal social support. This scale assesses the subjective perception of support regarding the sharing of emotions with close persons, and emotional help from family and friends (Slavin, Creedy, & Gamble, 2020; Zimet, Dahlem, Zimet, & Farley, 1988).

Working Alliance Inventory Short Revised (WAI-SR): the WAI-SR is a 12-item instrument designed to assess the quality of the therapeutic alliance (Hatcher & Gillaspy, 2006; Serralta, Benetti, Laskoski, & Abs, 2020). This scale measures the bond, agreement on therapeutic goals, and agreement on psychotherapy tasks, with each of these three domains having a separate score. The WAI-SR has both therapist-report and client-report (patient) versions, and both will be administered in this study.

System Usability Scale (SUS) and User Version of the Mobile Application Rating Scale (uMARS): the System Usability Scale (SUS) (Kortum & Sorber, 2015) and two sections of the User Version of the Mobile Application Rating Scale (uMARS) (Stoyanov et al., 2015) will also be used to evaluate the overall user experience regarding the use of Motherly.

Fidelity check: adherence to the single-session psychotherapy protocol will be assessed using a checklist adapted from a previous study on BA (Connolly Gibbons et al., 2023). Fidelity measurement will be conducted by an independent rater, rather than the psychotherapist, through audio recordings of the sessions. Fidelity will be defined by a total score representing the proportion of protocol items that were correctly implemented.

### Automated smartphone-based assessments

Patient-Reported Outcomes Measurement Information System (PROMIS): the PROMIS (Ader, 2007; Cella et al., 2010) (depression and anxiety, short form) will be available via the app (intervention group). Participants may respond at any time or via system notifications. These app-based scales are not part of the study outcomes; their function is to assist participants in symptom monitoring.

Participant interaction with the Motherly app will be recorded dynamically and continuously throughout the study. The app will record usage time, modules accessed, registered activities, and achievements, among other metrics. These records will be securely stored in a cloud server infrastructure without participant identification information, except for the study ID for subsequent linkage with the collected clinical data.

### Allocation concealment and masking

Most RCTs of psychological interventions report using masked outcome assessment, yet few describe in sufficient detail how masking was actually implemented and maintained (O’Keeffe, 2026). Since neither participants nor clinicians can be masked in psychotherapy trials, the estimates of the intervention effect rely on the integrity of outcome assessor masking. We describe below the strategies that will be adopted throughout the study to preserve concealment and masking.

The randomization list will be generated by an independent statistician and allocation will occur automatically via REDCap, ensuring concealment in both processes. Staff will be formally designated as masked or unmasked: outcome assessors will have no role in recruitment, allocation notification, intervention delivery, or fidelity assessment. To prevent unmasking during assessments, participants will be asked not to disclose any information related to group assignment. Measures that would reveal allocation (e.g., working alliance and app experience questionnaires) will be collected via an online form, after the clinician-rated assessment. Adverse events will be recorded separately by the study manager.

Access to allocation status and other group-related records in REDCap will be restricted through data access groups and role-based permissions. Because the study is conducted remotely, outcome assessors cannot be unmasked by incidentally overhearing group-related information. However, all staff will be instructed to confine treatment-related messages, e-mails, and case discussions to channels restricted to unmasked team members, and treatment-related matters will be discussed in a dedicated session with psychotherapists from which assessors will be excluded.

At the end of the post-treatment assessment, assessors will be asked to guess the participant’s allocation and rate their confidence. Success of masking will be quantified with an group-specific index based on assessors’ guesses and confidence ratings, reported descriptively with 95% confidence intervals to determine whether correct guesses exceeded chance. Suspected or confirmed unmasking will be registered and the affected participant reassigned to another assessor.

Finally, the main findings analysis (effect of the intervention on primary and secondary outcomes) will be conducted via a pre-specified R script using a dataset with masked group labels.

Table 5 specifies information concealment for each role involved, from researchers and participants to the statistical analyst.

**Table 5.** Masking information.

|  | Researcher | Management | Assessor | Clinician | Clinical supervisor | Participant | Analyst |
|---|---|---|---|---|---|---|---|
| Allocation | ● | ○ | ● | ○ | ○ | ○ | ● |
| Assessment | ● | ○ | ● | ● | ● | ○ | ● |
| Retention | ○ | ○ | ● | ● | ○ | ● | ● |
| Adherence | ○ | ○ | ● | ● | ○ | ● | ● |

*Legend*: ○=not masked; ●=masking; Researcher = project author (DF); Clinical supervisor = project author and clinical supervisor (PFZ); Management = study administration and logistics; Evaluator = assesses participants at baseline and outcomes; Clinician = delivers the intervention; Analyst = performs statistical data analysis at the end of the study.

### Adverse events

Adverse events (AE) are any unfavorable change or occurrence in a participant receiving an intervention, regardless of evidence of a causal relationship between the event and the intervention (Klein, Rozental, Sürig, & Moritz, 2024a). Despite extensive evidence that they are prevalent and deleterious (Strauss, Gawlytta, Schleu, & Frenzl, 2021), AEs are rarely assessed and reported in psychotherapy studies (Klein, Rozental, Sürig, & Moritz, 2024b). The present project will follow international (Guideline for Good Clinical Practice, 2025) and national guidelines and regulations (Brasil, 2020, 2024). Participants will be questioned regarding any unfavorable physical or mental health occurrence, whether related to the intervention or not.

Clinical worsening of depression will be considered an AE, defined as a 30% or greater increase in the QIDS-16 total score (clinician-rated [QIDS-C] or self-report [QIDS-SR]) at post-treatment (T3) or follow-up (T4) relative to baseline. This threshold is based on a previous validation study of the scale, which showed that the threshold for a clinically perceptible change is approximately 28.5% (Rush et al., 2003b). Additionally, we will assess serious adverse events (SAEs), which are defined here as: any adverse event resulting in death, life-threatening risk, situations requiring hospitalization or prolongation of hospitalization, significant disability, congenital anomaly, or a clinically significant event (Brasil, 2020). Suicide risk will also be considered a SAE and is defined here as a score of 2 or more on QIDS-16 Item 12 (Thoughts of Death or Suicide). This corresponds to endorsing either “I think of suicide or death several times a week for several minutes” or “I think of suicide or death several times a day in some detail, or I have made specific plans for suicide or have actually tried to take my life.”

All adverse events will be recorded, and in the case of a serious adverse event, the research ethics committee will be notified.

### Safety Considerations/Harm Prevention

The study protocol includes systematic procedures for identifying and managing safety concerns, particularly suicidal ideation or behavior, and domestic violence. While active suicidal ideation is an exclusion criterion assessed during recruitment, our protocol acknowledges that suicidal ideation or behavior may emerge during the study.

Identification and Assessment of Suicidal Risk: the identification of potential AEs and SAEs will be performed automatically in REDCap. If the system detects a 30% or greater increase in the QIDS-16 total score (QIDS-C or QIDS-SR), or a score of 2 or more on Item 12 of the instrument, an email notification will be sent immediately to the Principal Investigator and to the Management team. If during the session a participant indicates suicidal thoughts (e.g., “I wish I were not alive,” “Things would be easier if I were gone,” “I have thoughts of harming myself”), the therapist will conduct a direct, assessment of risk. If active suicidal ideation is confirmed, the standard intervention protocol (behavioral activation and app-based support) will be discontinued. Instead, the therapist will develop a written safety plan collaboratively with the participant. The safety plan documents: (1) warning signs and triggers for suicidal thoughts; (2) early coping strategies (relaxation techniques, physical activity, thought management); (3) people and social contacts to reach out to for support; (4) professional resources and crisis hotline numbers; and (5) reasons for living. The plan is shared in writing with the participant and stored in an accessible format (printed, screenshot, or other method agreed upon by participant and therapist). Following safety planning, participants with active suicidal ideation will be referred for immediate psychiatric evaluation and treatment. Participants will be contacted by a designated psychiatrist.

Domestic violence: any indication of domestic violence will be addressed with sensitivity and appropriate referrals to specialized services (women’s protective services, legal support, shelters, community resources). The therapist will assess available support networks and help the participant identify safe resources and action plans.

### Referral

During the study, participants will be referred for treatment in cases where depression progresses to a severe state, such as suicidal ideation, impairment that excessively compromises functionality, the emergence of psychotic symptoms, among others. At the end of the study period, participants who do not show remission of depressive symptoms will be referred to specialized mental health services within the Institute of Psychiatry of the Hospital das Clínicas or other specialized services geographically close to the participant.

### Sample size

Sample size was calculated based on the expected effect size of single-session psychotherapeutic interventions. For this purpose, data from the Metapsy platform (www.metapsy.org) were used, a research project led by Prof. Pim Cuijpers of Vrije Universiteit Amsterdam, which provides open data and tools for conducting meta-analyses of psychotherapy studies on various outcomes. Specifically, the “Depression: Psychotherapy vs. Control” database (version 24.0.2) (Miguel, Harrer, & Plessen, 2025) was used, which contains RCTs of psychotherapeutic treatments for depression published until December 2023. Filtering 900 RCTs for studies that tested single-session interventions for depression in adults, eight studies were found. Meta-analyzing these resulted in a standardized effect size of g=0.60 (95% CI 0.09, 1.11; p=0.027; I2=68%). Therefore, considering a type I error probability of 5%, statistical power of 80%, and a dropout rate of 15%, a sample size of 104 participants is required to detect an effect of 0.60. Sample size calculations were performed using G*Power software version 3.1.9.4 (Faul, Erdfelder, Lang, & Buchner, 2007).

### Pilot study

A pilot study will be conducted to test the study procedures and logistics. Based on feedback from coordinators, staff, and participants, adjustments to the study protocol will be made.

### Ethics

The study protocol was reviewed and approved by the Ethics Review Board of the Hospital das Clínicas da Faculdade de Medicina da Universidade de São Paulo (Comissão de Ética para Análise de Projetos de Pesquisa, CAPPesq; CAAE: 97532226.8.0000.0068). Written informed consent will be obtained from all participants prior to their inclusion in the study. Our trial will be conducted in accordance with the Declaration of Helsinki.

### Data analysis

Generalized estimating equations (GEE) models will be used to examine the effects of the intervention on both primary and secondary outcomes. GEE is a semiparametric approach that accounts for within-participant correlation across repeated assessments and yields valid inferences without requiring correct specification of the correlation structure (Twisk, 2013). Because repeated observations are expected to be correlated over time, an autoregressive order-1 (AR1) correlation structure will be used in all models. All statistical inference on GEE coefficients will be based on a robust sandwich variance estimator.

Analyses will be conducted in the intention-to-treat (ITT) sample, and therefore all randomized participants will be included in the analyses. Missing data will be handled using multiple imputation by chained equations (MICE) (Azur, Stuart, Frangakis, & Leaf, 2011; Rubin, 2004), with data assumed to be missing at random (Jakobsen, Gluud, Wetterslev, & Winkel, 2017). One hundred imputed datasets will be generated. Each model will be fitted separately in each imputed dataset, and the estimates will be pooled using Rubin’s rules. The fraction of missing information (FMI) will be reported for each pooled parameter, and values above 0.7 will be flagged (Graham, Olchowski, & Gilreath, 2007).

Following the strategy outlined by Twisk et al (2018), models will include outcomes as dependent variables, time as categorical variables (with baseline, T1, as the reference group), and the time-by-group interaction, with the formula:

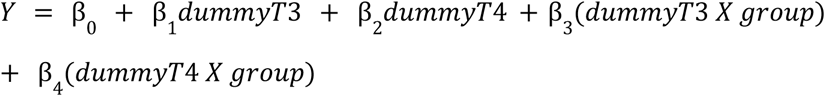

Where *Y* is the outcome, *dummyT3* and *dummyT4* are dummy variables for the post-treatment (week 4) and follow-up (week 8) assessments, and *group* indicates allocation to the intervention (1) or waitlist (0) group. Groups are assumed equal at baseline by randomization, so the intercept (β_0_) represents the common baseline mean. β_3_ and β_4_ estimate the effect of the intervention at T3 and T4, respectively, which is analogous to adjusting for the baseline value of the outcome. For the primary outcome (QIDS-C16), the model includes T1 and T3 only, so β_3_ is the primary estimate of the intervention effect. Secondary outcomes will be modeled at T1, T3 and T4. The Clinical Global Impression – Improvement (CGI-I) is assessed only at T3 and has no baseline value. It will therefore be analyzed as a continuous score (1–7) using linear regression on group, with the same imputation and pooling procedures.

Estimated marginal means will be obtained for each group and timepoint. Between-group contrasts (waitlist − intervention) will be computed at each follow-up assessment. Standardized effect sizes (Cohen’s *d*) (Cumming, 2011), with 95% confidence intervals, will be calculated for each contrast. Each *d* will be the between-group difference in marginal means divided by the pooled within-group standard deviation at that timepoint, averaged across the imputed datasets.

A sensitivity analysis will assess the influence of missing data on the results. The same models will be fitted without imputation in a complete-case sample, which for each outcome includes only participants with observed data at every timepoint analyzed for that outcome. Analyses will also be performed to verify the association of baseline characteristics with dropout (participants who left the study or did not attend the post-T1 assessments). All statistical tests will be two-sided with a significance criterion of *p* < 0.05, and 95% confidence intervals will be reported for all estimates. Analyses will be conducted in R (version 4.4.3), using the geepack (GEE), mice (multiple imputation) and emmeans (marginal means, contrasts and effect sizes) packages. As mentioned before, they will follow a pre-specified R script applied to a dataset with masked group labels. (https://github.com/dfatori/motherly-plus).

### Data management

Data from forms and interviews will be collected via REDCap and stored on the servers of the Núcleo Especializado em Tecnologia da Informação (NETI, Specialized Information Technology Center) at the Hospital das Clínicas Faculdade de Medicina da USP, which follow quality and security standards and provide automated backups. Specific information that could identify a particular individual (name, telephone, or physical and electronic addresses, etc.) will not be included in the databases. The identification of each participant will be conducted solely through a numerical code. Data will be stored on secure servers (Google Drive, Motherly app proprietary servers, HC-FMUSP REDCap servers), to which only the candidate, the group’s research manager, and the coordinators will have access. Data collected via the app will be stored on Digital Ocean servers, also following quality and security criteria. Data extracted from both REDCap and Digital Ocean will be anonymized and stored on an institutional Google Drive for data analysis. Audio recordings of psychotherapy sessions will be stored in a compressed and encrypted format, only to be opened for the evaluation of intervention fidelity. Regarding data sharing, after the publication of main findings, data will be available upon request.

### Trial management

This trial will be overseen by three bodies: a Trial Management Committee, a Trial Steering Committee, and the Ethics Review Board of the Hospital das Clínicas da Faculdade de Medicina da Universidade de São Paulo (Comissão de Ética para Análise de Projetos de Pesquisa, CAPPesq). The Trial Management Committee is composed of the principal investigators (DF, PFZ) and trial managers, who will meet weekly to coordinate and monitor the progress of the trial and to ensure that the protocol is adhered to. This committee will also be responsible for monitoring any adverse event and reporting it to the other committees (and taking action to safeguard participants), as well as for ensuring the quality of the trial.

The Trial Steering Committee is composed of senior researchers from our group (DF, PFZ and will meet once a month to oversee the trial, ensure that it is conducted according to the protocol, and monitor its progress regarding recruitment and retention of participants, the occurrence of adverse events and actions needed to safeguard participants, and adherence to the study timeline. This committee will also be responsible for decisions about the continuation, modification, or termination of the study. Any modifications to the trial will only be implemented after approval by CAPPesq, in which case funders will be notified and the protocol will be updated in the clinical trial registry (ClinicalTrials.gov). The Trial Steering Committee will also meet with funders every six months to report on the status of the trial.

CAPPesq is responsible for reviewing all procedures involved in this trial, assessing its relative risks, and determining what action should be taken to safeguard participants. This board reviewed and approved the study protocol and informed consent form prior to data collection (CAAE: 97532226.8.0000.0068). Since this intervention is considered low risk, no periodic meetings with CAPPesq are planned; however, additional meetings may be scheduled if a serious adverse event occurs or if the Trial Steering Committee decides to amend the trial protocol. Given that the intervention was rated as low risk, no Data Monitoring Committee was considered necessary.

### Funding

This project is funded by (1) the Fundação de Amparo à Pesquisa do Estado de São Paulo (FAPESP; Process: 21/12901-9) through the Centro Nacional de Ciência e Inovação em Saúde Mental (CISM); and Conselho Nacional de Desenvolvimento Científico e Tecnológico (CNPq; Process: 408132/2025-6). The funders had no role in study design, nor will have in data collection, data analysis, data interpretation, or writing of the paper. The authors declare no conflict of interest.

### Dissemination policy

Results of this trial will be published as a preprint and submitted for publication in peer-reviewed journals.

## Discussion

We described the design and methods of a parallel-group RCT evaluating the efficacy of a smartphone-assisted single-session psychotherapy intervention for PPD. To our knowledge, our study represents the first RCT incorporating digital support into this approach. Beyond evaluating efficacy across maternal symptoms, our study is also designed to investigate moderators, mediators, and treatment-specific processes, including therapeutic alliance and app engagement.

The design of the study has several strengths. First, we will implement masked assessments of the primary outcome with a clinician-rated scale, a methodological improvement relative to RCTs for PPD. The majority of trials in this field rely on the EPDS as their outcome measure (Hua, Yue, Zhu, Yang, & Zhou, 2025), which has items assessing not only depression but also symptoms of anxiety, and deliberately omits appetite, sleep and weight items (Matsumura et al., 2020). The QIDS-C16 covers all nine DSM-5-TR symptom criteria for major depressive disorder and is administered by a trained clinician who probes the presence and severity of each symptom. Furthermore, masked administration is important in a waitlist-controlled design, in which participants are necessarily aware of their allocation, and self-reported symptom change may be influenced by expectation in the intervention group and by disappointment in the control group (Sterne et al., 2019b). Together with an independently generated randomization list, automated allocation through REDCap, and a masked analyst, the use of clinician-rated scales addresses bias that currently limit single-session therapy studies, in which trials have been rated as high risk of bias (Kim, Ryu, & Chibanda, 2023b).

Treatment fidelity will be rated by an independent assessor from audio recordings of all sessions. This type of assessment is scarce in psychotherapy research, with evidence that only 10.7% of studies implement adequate procedures to check integrity of the intervention (Cox, Martinez, & Southam-Gerow, 2019). Additionally, the treatment manual will be publicly available. These features are intended to facilitate replication and produce a low-risk-of-bias estimate of efficacy that can serve as a benchmark for subsequent trials using active controls.

Another strength of the study lies in the assessment of adverse events. Despite extensive evidence that adverse events are prevalent and deleterious (Strauss et al., 2021), this practice is rarely done in psychotherapy trials (Klein et al., 2024a). Finally, due to its brevity, the intervention controls for the cumulative effect of several psychotherapy sessions. Therefore, the single session design of the study is adequate to explore the mechanisms through which the intervention works.

Despite the strengths of the study, it will face several challenges. The recruitment strategy using social media, and the need for internet access and a smartphone might result in a sample with mean income higher than the average of the Brazilian population (Pedro F. Zuccolo et al., 2024). This feature might hinder women from the lowest socioeconomic strata, who are in most need, to participate, and might select women with greater digital access and literacy. Moreover, the use of wait-list can inflate the estimate of the effects of the intervention (Gold et al., 2017b), and can produce nocebo effects (Cunningham, Kypri, & McCambridge, 2013). However, the brevity of the intervention might mitigate these effects, and participants will be monitored throughout the study for safety.

The delivery of the intervention in a single session may also present some challenges. Since single session therapy is not the hegemonic model of psychotherapy, it could be challenging to change clinicians’ mindset, who are used to applying psychotherapy protocols with more than one session. Moreover, single session psychotherapy does not allow for course correction as therapists will see the patient only once. Therefore, our study might require extensive training and more supervision than anticipated to tackle these obstacles.

As a final issue, what is being tested is a package that consists of both the single session therapy and the use of the app. Though we have metrics to assess the association between app usage and symptom reduction, it will not be possible to distinguish which component is most responsible for the effect.

This RCT to test the efficacy of a smartphone-assisted single-session psychotherapy intervention for PPD has potential public health relevance. By testing an intervention package that combines single-session psychotherapy and technology, there is high potential for addressing the mental health gap and growing burden of PPD (Fusar-Poli et al., 2025; Patel et al., 2018). Because the session is delivered remotely and subsequent support is provided through a smartphone app, the intervention does not require participants to travel to a clinic and can reach participants in remote and underserved areas. The brief, single-session format also makes more efficient use of a scarce specialized workforce, an important consideration in settings where mental health professionals are unevenly distributed. Results from this first trial will inform improvements on the intervention and designing of futures tests with active controls.

## Supporting information

SPIRIT checklist

## Data Availability

All data produced in the present study are available upon reasonable request to the authors

