## Supplementary material for "Motherly Plus: Study Protocol of a Randomized Controlled Trial Evaluating a Smartphone-Assisted Single-Session Psychotherapy Intervention for Postpartum Depression"

Pedro F Zuccolo, Henrique Melo, Leonardo Seda, Tatiane Borja, Eduarda Souza, Julia Nicolella, Leonardo Seda, Alicia Matijasevich, Wagner Ribeiro, Renatha El Rafihi Ferreira, Daniel Fatori

**https://www.medrxiv.org/**

Intervention manual

**Intervention Protocol Guide**

### 1. Introduction

This intervention consists of a single-session psychotherapy protocol based primarily on behavioral activation (BA), followed by a period in which participants use a smartphone application developed to treat maternal depression and other mental health symptoms (Motherly app). It was designed to promote, in a scalable way, two of the basic mechanisms thought to underlie the reduction in depressive symptoms produced by BA: increased contact with positive reinforcers and a reduction in the avoidance behaviors that limit access to reinforcing activities. The single therapy session focuses on three components: psychoeducation about the behavioral model of depression, collaborative problem identification and targeting, and training in how to use the app to implement these strategies over the following weeks.

#### *1.1. Theoretical foundations*

Behavioral activation (BA) is a brief, structured psychotherapy modality with decades of evidence demonstrating its efficacy in treating depression (Abreu, 2006; Dimidjian, Barrera, Martell, Munõz, & Lewinsohn, 2011; Kanter et al., 2010; C. W. Lejuez, Hopko, Acierno, Daughters, & Pagoto, 2011) and with demonstrated equivalence to more complex psychotherapeutic interventions (Richards et al., 08/2016) and to medication (Moradveisi, Huibers, Renner, Arasteh, & Arntz, 2013). Its simplicity has already allowed for adaptations to be delivered by non-specialists (Ekers, Godfrey, et al., 2011; Ekers, Richards, McMillan, Bland, & Gilbody, 2011; Singla et al., 2025), via the internet, and through computers and smartphones (Huguet et al., 2016).

BA is based on a behavioral theoretical model (Dimidjian et al., 2011; Lewinsohn PM, Hoberman H , Teri L, Hautzinger M, 1985; Martell, Dimidjian, & Herman-Dunn, 2013), according to which depression involves a maladaptive interaction pattern between the individual and their environment maintained by two processes: the absence of contact with positive reinforcement contingencies and avoidance behaviors(Martell et al., 2013). A positive reinforcement contingency is understood as a condition in which an action is followed, contingently, by the occurrence of a reinforcing stimulus (Catania, 1999). Reinforcing stimulus is an event that increases the future probability of that action occurring under similar conditions (C. B. Ferster & Skinner, 1957). In a clinical context, positive reinforcers are consequences generally associated with a sense of well-being or pleasure, and a feeling of accomplishment or efficacy, such as social approval, a sense of competence, pleasure, mastery, or progress toward personal goals. According to the BA model, depressive episodes involve a chronic reduction in response-contingent positive reinforcement (Lewinsohn PM, Hoberman H , Teri L, Hautzinger M, 1985), which can occur either because reinforcement is unavailable (e.g., relationship breakup, job loss, bereavement, etc.), because the person is less sensitive to it, or because of a pattern of avoidance and passivity that reduces access to reinforcement. Avoidance behaviors are understood as actions that are strengthened or sustained because they result in the removal or postponement of a stimulus or situation. In a clinical context, this translates into delay or procrastination when facing tasks, decisions, interactions, or contexts that may bring discomfort or the risk of failure (Charles B. Ferster, 1973). Avoidance behaviors produce momentary relief from discomfort and are thereby strengthened, but in the medium and long term they result in a more restrictive, inflexible, and passive behavioral repertoire, which prevents contact with situations involving important positive reinforcers.

Based on the behavioral conception of depression, BA aims to increase contact with positive reinforcement contingencies and decrease avoidance and passivity behaviors (Dimidjian et al., 2011). This translates into practices intended to engage the individual in behaviors and situations that may be experienced as pleasurable or productive, that provide a sense of belonging, and that move the person toward a purpose that matters in their life history (Dimidjian et al., 2011). By engaging in these activities, the individual can gradually acknowledge and soothe challenging emotions and thoughts while promoting more functional ones. Therefore, the BA session focuses on action and problem-solving, which requires testing new ways of perceiving and behaving in everyday situations, guided by a plan rather than exclusively by momentary mood or motivation. Furthermore, BA recommends helping the person avoid behaviors that involve isolation or that may strengthen challenging emotions.

The primary goal of BA is to reduce depressive symptoms using three main strategies: (a) engagement in positively reinforced behaviors, which involves situations and activities that promote a sense of accomplishment, pleasure, or belonging; (b) the reduction of isolation and avoidance behaviors; and (c) the development of problem-solving skills. It is an intervention focused on practical action: the therapist helps the person find new ways of behaving in, and relating to, the everyday situations that matter to them, across different life domains (leisure, productivity, and relationships).

Over the past two decades, the principles of BA have been consolidated into structured, manualized protocols. Martell and colleagues (Martell et al., 2013) organized the accumulated evidence and clinical techniques into a comprehensive treatment package in which behavioral avoidance is the primary target and clinical decisions are guided by an idiographic functional analysis. In this framework, assessment involves identifying target behaviors, the consequences that maintain them, and the environmental contexts in which they occur (antecedents), so as to map the sources of reinforcement sustaining depressive behavior and the contingencies that could strengthen alternative, healthy behaviors. The protocol also incorporates strategies more indirectly related to activation, such as addressing rumination, mindfulness training, and self-reinforcement. This comprehensiveness makes it particularly suited to more complex clinical presentations, but the level of assessment it requires demands substantial time and specialized training, which limits its feasibility in brief intervention contexts (Carl W. Lejuez, Hopko, & Hopko, 2001).

Lejuez and colleagues (Carl W. Lejuez et al., 2001; C. W. Lejuez et al., 2011) addressed this limitation by developing the Brief Behavioral Activation Treatment for Depression (BATD, later revised as BATD-R), a compact protocol restricted to the components most directly tied to activation (e.g., activity monitoring and scheduling), organized within an idiographic, values-driven framework. A comprehensive functional analysis is not conducted; instead, potential sources of reinforcement are identified indirectly, through the participant's values across multiple life areas. Values are defined as broad descriptions of how a person would like to live within a given life area, and activities are their most concrete and reducible manifestation: specific behaviors that can be performed on a daily basis, within the person's current abilities and resources. The link between values and activities is precisely what preserves the behavioral rationale of the approach. Because depressive behavior is largely maintained by negative reinforcement that is immediate and certain, the activities selected to compete with it must yield reinforcement that is also immediate and salient. Activities anchored in values are more likely to be experienced as enjoyable or meaningful and to be sustained over time, thereby increasing the density of positive reinforcement in the person's repertoire and progressively displacing depressive behavior. Considering several life areas, rather than only one or two, further broadens the opportunities for reinforcement and for success experiences. This streamlined structure has also proven to be dose-flexible: significant reductions in depressive symptoms have been reported with six to eight sessions, and even with a single session (Gawrysiak, Nicholas, & Hopko, 2009) .

Our group has previously adapted this brief rationale to the Brazilian context, delivering a four-session, BA-based protocol online to 81 pregnant women with depressive symptoms, supported by the Motherly app (Fatori, Zuccolo, Xavier, Matijasevich, & Polanczyk, 2023; Zuccolo, Xavier, Matijasevich, Polanczyk, & Fatori, 12/2021). The intervention proved feasible and acceptable in a nationwide, fully remote format. The present protocol extends this progressive simplification one step further, condensing the therapist-delivered component into a single session followed by four weeks of app-supported activity planning and monitoring, and organizing activities across three life domains: leisure, productivity, and relationships.

*1.2. Overview of the intervention protocol*

The intervention provided to participants will consist of three distinct phases, as outlined in the table below:

| **Protocol general structure** |
| --- |
| 1. **Pre-session questionnaire:** The participant must complete this questionnaire before the session. It asks about her current and past sources of reinforcement and about her support networks. The information collected will guide the session, so that the intervention can be tailored to her specific needs. |
| 2. **Psychotherapy session:** The session lasts 90 minutes. You will begin by establishing rapport with the participant and providing a safe, supportive environment. You will then gather information about relevant areas of her life, such as her support networks and current daily routine. The clinical work is based on Behavioral Activation, a therapeutic strategy described in detail in section 3.1.2. In the final part of the session, you will introduce the Motherly app, show her how to use it, and encourage her to engage with it. |
| 3. **Motherly app:** After the session, the participant will use the app for the subsequent weeks. Motherly provides ongoing support for the intervention: she can schedule behavioral activation activities, monitor her symptoms over time, and access psychoeducational content organized into CBT-based modules such as emotional regulation, sleep hygiene, and mindfulness . |

*1.2.1. The pre-session questionnaire*

The questionnaire marks the beginning of the treatment process. Because you will only have a single session with the participant, it is essential that she arrives at the appointment having already reflected on some key questions. This allows the session time to be used more efficiently. Completing the questionnaire begins the process of reflection on the issues to be addressed in psychotherapy. During the initial contact, you will introduce yourself and ask her to complete the questionnaire. After 24 hours, you will check on REDCap whether she has completed it and then schedule the session.

The questionnaire is formatted as follows:

| |
|---|
| ***"Before we schedule your psychotherapy session, we would like you to answer a few questions. Please reflect on them and try to answer with care. These questions mark the beginning of your therapeutic process. Answering them will help you get the most out of your session and will assist your therapist in guiding you. There is no need to write long answers."*** <br> *What does a typical day look like for you, from morning to night? What do you usually do?* <br><br> *What do you enjoy or used to enjoy doing during your leisure time to have fun, relax, or feel good about yourself?* <br><br> *What routine responsibilities, work tasks, household chores, or self-care activities do you currently carry out or feel you should be doing?* <br><br> *Are there activities you used to do (for pleasure, obligation, or habit) that you stopped doing or are doing less frequently since your symptoms began (or since becoming a mother)?* <br><br> *Which people (family members, friends, etc.) do you enjoy spending time with or can ask for help when needed?* <br><br> ***"Thank you very much for taking the time to share your answers. They will be carefully reviewed by the psychotherapist and thoughtfully addressed with you during your session."*** |

*1.2.2. Structure of the single-session psychotherapy protocol*

The therapy session comprises several distinct stages that must be completed. Below is a summary overview of each stage:

**Part I – Introduction:** In the initial phase, you will introduce yourself and provide psychoeducation on how the session will work. After that, there will be a period during which the participant can speak freely about her chief complaint, the difficulties she has been facing with depression and motherhood, and any other topics she wishes to raise. During this phase, you will provide a supportive environment so that the participant feels comfortable, while asking targeted questions to gather information that helps you understand her current difficulties.

**Part II – Assessment:** You will guide the conversation to gather information on specific domains of the participant's life. Here, you will discuss her current daily routine, existing sources of reinforcement, support networks, and prior experiences. The information gathered in this section will serve as the foundation for Part III (Behavioral Activation).

**Part III – Behavioral Activation:** This portion of the session is dedicated to Behavioral Activation (BA). First, explain the rationale for BA in clear, accessible language attuned to her daily life, without relying on technical jargon. Next, you will work with her to identify activities that are relevant and meaningful to her. Finally, help the participant select and schedule specific BA activities to be carried out over the coming weeks.

**Part IV – The Motherly App:** Once the behavioral activation activities have been defined, you will introduce the Motherly app. You will guide the participant through installing it and scheduling the agreed-upon activities for the coming weeks. You will then present the remaining modules available in the app. If a particular module seems especially relevant to that participant, based on what came up in the session, explore it with her and encourage her to use it.

**Part V – Closing:** In the final section of the session, you will review the treatment framework with the participant and summarize what was accomplished. At this stage, you may also ask what she is taking away from the session, whether she has any remaining questions, and whether she has any feedback. Before concluding, briefly reinforce the commitments made for the coming weeks and emphasize the importance of trying to put them into practice despite any obstacles that may arise.

*1.2.3. Motherly app*

The Motherly platform consists of a suite of modules comprising specific activities (described below). The intervention is grounded in three core concepts: psychoeducation, behavioral and activity monitoring, and gamification elements.

Participants receive psychoeducational information regarding the importance of health-promoting behaviors and maternal and child healthcare, taking into account their impact on the future development of their children. Additionally, scheduling features and automated notifications are provided to assist in organizing and tracking daily activities, thereby contributing to the reduction of depressive symptoms. Gamification elements play a critical role in sustaining user engagement and fostering motivation toward healthy behaviors. Gamification is defined as the application of game-design elements in non-game contexts to engage users and facilitate problem-solving.

The app is structured into five primary domains: Home, Library, Health, Pregnancy, and Profile, organized to address distinct dimensions of maternal care, including mental health, sleep hygiene, nutrition, physical activity, social support networks, pre- and postnatal care, and health education resources. Within the “Home” section, mothers can log and monitor appointments, as well as view tailored content recommendations. The “Library” tab provides essential information relevant to the prenatal, perinatal, and postnatal periods. The “Health” section delivers content focused on mental health promotion, including interactive features to implement behavioral activation strategies. In the “Pregnancy” section, users can track gestational progress and infant developmental milestones. Finally, the “Profile” section allows users to monitor their overall well-being and track achievements and progress throughout their use of the app.

The Motherly app is available on the Play Store as internal testing so that only participants can download it. The research team has also recorded an introductory video detailing the app’s primary features and user interface, available at https://youtu.be/f2IOSdgHo64.

This material provides a brief overview of the platform and its features; it does not, however, replace exploring the software yourself. We recommend navigating Motherly on your own to become more familiar with what it offers and to form your own impression of how it works.

**2. Postpartum Depression (PPD)**

All study participants have a diagnosis of PPD, as this serves as an inclusion criterion for the research. Table 1 below presents the primary signs and symptoms to assist the therapist in identifying potential participant difficulties. From a clinical perspective, certain PPD symptoms overlap with those observed in Major Depressive Disorder. This material is intended to support clinical listening and the observation of core maternal factors throughout the session.

**Table 1.** Moods, Thoughts, and Physical Health/Behaviors Associated with Maternal Depression.

| Mood | Thought | Physical Health / Behaviors |
|---|---|---|
| • Loss of interest or pleasure (anhedonia)<br>• Feelings of guilt<br>• Mood swings<br>• Anxiety<br>• Hopelessness<br>• Fear of being alone with the baby due to coping difficulties | • Difficulty concentrating<br>• Low self-esteem<br>• Loss of focus<br>• Difficulties in decision-making and thought processes<br>• Suicidal ideation<br>• Excessive worry<br>• Irritability<br>• Self-doubt regarding activities such as caring for the baby | • Loss of appetite<br>• Changes in physical appearance<br>• Sleep disturbances<br>• Frequent crying<br>• Low energy |

*Source:* Adapted from *Registered Nurses' Association of Ontario* (2018), based on Banti et al. (2009), O'Hara & McCabe (2013), and Zauderer & Davis (2012).

PPD presents specific clinical manifestations. In this context, mothers commonly develop negative perceptions of their parenting competence, accompanied by reduced confidence in their parental abilities. This process can lead to a decrease in maternal self-efficacy, defined as a mother's belief in her capacity to adequately care for and respond to her infant's needs. Furthermore, PPD is associated with diminished maternal sensitivity, which refers to the ability to accurately perceive and interpret the infant's cues and respond to them appropriately.

Another frequently observed feature in mothers with PPD is elevated parental stress, which refers to psychological distress resulting from the stressors inherent in the demands and responsibilities of child-rearing. Generally, this stress is experienced as a negative or aversive emotional response to parenting tasks. High levels of parental stress may be associated with dysfunctional, less effective, and inconsistent parenting practices, as well as negatively impacting maternal sensitivity. Consequently, mothers with PPD tend to describe their infants as more difficult to care for, attributing a difficult temperament to them, interpreting their crying as excessive, and perceiving them as more demanding, which contributes to a more dysfunctional mother-infant interaction.

Given this context, mothers with PPD tend to display a lower frequency of positive interactions with their infants. Compared to non-depressed mothers, they may appear less engaged, exhibiting reduced physical touch, vocalization, and eye contact with the child. Additionally, they may express heightened negative affect and demonstrate less spontaneity during interactions. In some cases, a reduction in affective expression is observed alongside irritability, aggressive impulses, or even feelings of rejection toward the infant. Mothers also tend to participate less frequently in interactive activities with the child—such as reading, games, and play—which are critical factors for strengthening the mother-infant bond and, consequently, for the child's socioemotional development.

Figure 1 illustrates the maintenance cycle of characteristic PPD symptoms and their impact on mother-child interaction.

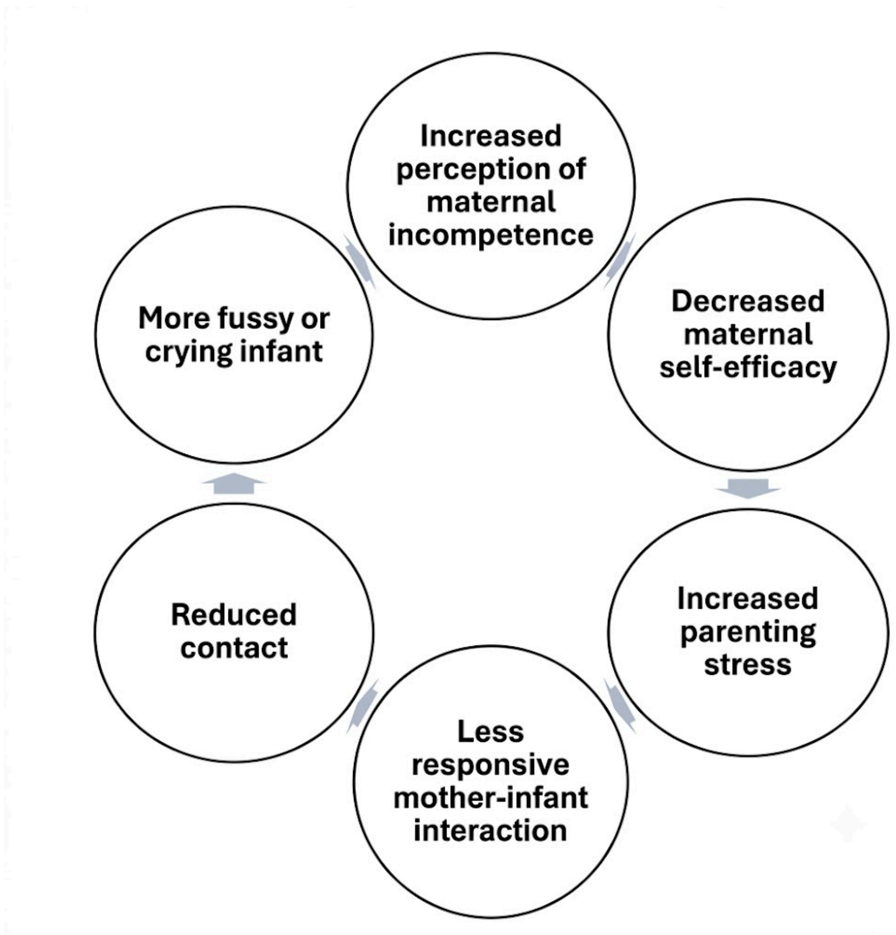

**Figure 1.** Maintenance cycle of Postpartum Depression symptoms.

### 3. About single-session psychotherapy

As a therapist, you have likely had clinical experiences in which psychotherapy was conducted over multiple sessions, often without a predetermined number of meetings. In this format, the initial sessions typically aim to get to know the participant, build a therapeutic alliance, and formulate a clinical assessment. Often, the interventions deemed appropriate for the case are implemented only after this initial diagnosis. The focus of this study is to work with a different form of therapy. The idea is to develop a short-term intervention (single-session). For this to work, certain points need to be kept in mind. Based on single-session literature (Dryden, 2021; Hoyt, 2024) and the study's rationale, the following aspects are listed to assist in understanding this intervention proposal:

#### 3.1. A growing field of research

Since the 1990s, single-session psychotherapy has increasingly become the subject of studies worldwide. Considering that interventions planned to occur over many sessions tend to present higher attrition rates (Fernandez, Salem, Swift, & Ramtahal, 2015), and that longer treatments do not always result in better outcomes in psychotherapy (Ciharova et al., 2024), brief intervention formats have gained traction as potentially effective alternatives in mental health. In this context, a recent meta-analysis (Schleider et al., 2025), which aggregated results from 24 literature reviews, showed that the majority of them (24 reviews) found positive effects for single-session psychotherapy. This finding reinforces the potential of this modality of psychotherapeutic intervention. We now seek to advance further in this field by studying its effects on the treatment of maternal depression.

#### *3.2. Clinician expectations vs. Participant needs*

Generally, in both research contexts and private practice, a significant percentage of participants do not continue therapy for the expected number of sessions. A portion of these participants, in fact, attend only one session and do not return. According to a traditional or common view of psychotherapy, these cases are often seen as failures, dropouts, etc. However, research indicates that, in reality, many participants report improvement even when attending a reduced number of sessions. Some report feeling significantly helped in one or a few sessions. Not all participants seeking therapy wish to continue a long-term process. Research conducted with participants who attend only one session suggests meaningful improvements in their mental health symptoms and in their satisfaction with the care received (Talmon, 1990).

*3.3. Importance of diversifying modalities of care in psychotherapy*

Despite evidence that psychotherapy is effective for a range of mental disorders, there are several barriers to accessing this type of treatment. A large portion of the Brazilian population does not have the financial resources to afford long-term, weekly psychotherapeutic treatment. In addition, few psychology professionals are available in the public health system relative to the existing demand. Thus, it is of great importance to develop mental health care modalities that are more scalable and accessible. Researching and developing shorter forms of care supported by digital technologies is one way to advance and democratize psychological treatment.

*3.4. Clinical perspective for single-session*

The clinical perspective involved in working with a single session differs, in several respects, from the one usually adopted in long-term psychotherapy services. A few guiding principles are fundamental here and should shape how the single session is conducted.

| |
|---|
| **The time you have is this session**<br>This is a central aspect of single-session psychotherapy. The time you and the participant have to work on the issues identified is this single session. Stay focused and strive to use the time wisely. |
| **Work with existing resources**<br>Even though the participant may find herself in a fragile or vulnerable state, she possesses her own resources—such as life experiences, personal skills, strengths, and support networks—that can be drawn upon to build alternatives. Similarly, as a therapist, you bring experience, knowledge, clinical training, and peer consultations, all of which are of fundamental importance. Harnessing the resources of these two primary actors makes it possible to conduct productive therapeutic work. |
| **Focused help can be of great value**<br>Just because psychotherapy is delivered in a single session does not mean you should pressure yourself to achieve a major breakthrough or a highly complex intervention that will yield radical changes in the participant's life. If the participant leaves with even one outcome she considers useful, that holds great value. A single, meaningful insight or |

| |
|---|
| strategy is far better than leaving with the impression of having been “bombarded” with a chaotic mix of methods, techniques, interpretations, and the like. |
| **Provide help in the here and now**<br>For therapists accustomed to routine clinical practice, it can be tempting to try to establish a complete case conceptualization or diagnostic formulation before intervening. A core premise of single-session psychotherapy is that you can help the participant even without knowing her full life history in exhaustive detail. Relevant past experiences will surface during the session to the degree that is necessary. |
| **Do not rush!**<br>Single-session work is targeted and bounded by a single encounter. Although this format can create a sense of urgency to resolve everything within the available timeframe, empirical literature on single-session psychotherapy indicates that this time is sufficient to complete the proposed work. Later in this manual, strategies for keeping the participant focused will be presented; however, it is crucial to avoid projecting a sense of time scarcity or making the participant feel rushed. |

### 4. Step-by-step session guide

The different stages of the session are described below, together with the rationale for each one and examples of how to work through it. You should try to complete all of these stages during the appointment whenever possible. The sample statements are illustrative. You can and should adapt your phrasing to your own interpersonal style, taking the participant's characteristics into account. You may also go into more depth in particular sections, depending on the participant's individual profile.

#### 4.1. Part I – Introduction

##### *4.1.1 Therapist's introduction*

Initial contact with the participant has already been made via WhatsApp; briefly reconnect with the participant by stating your name and credentials (e.g., "I am [Name], the psychologist who scheduled this appointment with you, and I will be conducting your session today").

##### *4.1.2 Psychoeducation on session dynamics*

Explain to the participant how the intervention works and how the session will be structured. Make it clear that your contact with her will be limited to this single session. Tell her that you will first talk about her life in a more open-ended way, and that you will then work together on strategies to address the difficulties she is facing. After that, you will show her how the Motherly app works; it will be the tool she uses over the following weeks to continue the work begun in the session.

*"Today, we will be working with a single-session brief psychotherapy format. We have 1 hour and 30 minutes to calmly discuss what is happening in your life and to collaboratively build strategies that are meaningful to you. Most importantly, these strategies are intended for you to continue using after our conversation, as day-to-day practice is where meaningful change begins.*

*To make optimal use of our time, I will explain how our conversation will be organized. We will have four main phases today: (1) first, I would like to hear about how you are feeling and the difficulties you have been experiencing; (2) next, I will ask you a few questions about specific areas of your life to better understand how to support you. You completed a pre-session questionnaire, and we will review the information you provided in that form during this part; (3) after that, I will work with you on a therapeutic strategy that may be helpful for your current situation; and (4) finally, I will introduce an app developed to support mothers experiencing postpartum depression. The goal is for you to install this app on your smartphone and use it to put the strategies we discussed into practice in your daily routine. Okay? Do you have any questions?*

*To ensure we make the most of our time, we will need to stay focused on our objectives for today. For this reason, I may need to interrupt you at certain points. This is a standard part of the process. That being said, there is no rush—we have sufficient time to talk at a steady pace. Is it alright if I interrupt you when necessary?"*

##### *4.1.3 Empathy and rapport building*

***"Well, I would like to learn a bit about how you are doing and what motivated you to participate in this research study."***

Allow the participant to share her reasons for joining the study and what has been happening in her life. Use this moment to understand her living situation, sociodemographic characteristics, and support networks. When you open the floor to her, create an environment in which she feels welcomed, respected, and comfortable expressing herself. During this stage, utilize active listening. Active listening is defined here in accordance with Rogers' conceptualization, as a way of listening to another individual to understand the meaning of their lived experience from their own frame of reference (Rogers & Farson, 1957). Operationally, this entails: a) listening without interrupting, judging, or making immediate interpretations; b) observing both verbal content and emotional tone, pauses, and speech intensity; c) identifying central feelings and main themes in the participant's narrative; d) reflecting back to the participant, in clear and simple language, what was perceived—namely, the verbal content, emotional tone, core feelings, and main themes. This can be accomplished, for example, by summarizing or paraphrasing her statements.

| |
|---|
| **Active listening:**<br><br><u>Paraphrasing</u> → consists of repeating what the participant said using fewer words.<br><br>Participant: *Lately I've been having relationship difficulties with my husband… ever since he brought up divorce, I've felt distant from him… (pauses, appears reflective)*<br><br>Psychotherapist: *(In a calm tone, looking gently at the participant) Hmm, he actually brought up divorce, I understand…*<br><br>Participant: *Yes… we used to get along so well. But lately, it seems like everything has become a reason for us to drift apart. (...)* |
| **Active listening:**<br><br><u>Summarizing/Rephrasing</u> → consists of expressing, in your own words, the most important aspects of what the participant said.<br><br>Participant*: And then I found out my husband lied to me, saying he had to work late. I am so angry, but at the same time, I don't know if I should talk to him right now… our relationship is already so strained, and I don't want to be the one who is always confronting him.*<br><br>Psychotherapist: *So even though you are angry, you hesitate to talk to him… as if the risk of drifting even further apart feels too high. You must be really worried about that.*<br><br>Participant: *Yes, I've been thinking about this a lot lately… I really want to keep things good between us, but I'm increasingly losing hope that things will improve. (...)* |

This stage is fundamental in single-session work and serves several key functions. First, it helps refine communication between therapist and participant. Upon hearing her words paraphrased or summarized, the participant receives a clear signal that the therapist is following her narrative closely. Furthermore, this process allows for the immediate clarification of any misunderstandings, increasing the likelihood that the participant feels genuinely understood and supported. Active listening also keeps the interaction flowing, helping the participant carry on with her account. For the therapist, repeating or reformulating what the participant has said deepens their own understanding of what is being shared. In doing so, the therapist may notice subtle details previously overlooked, or gain clinical insight into the material just paraphrased.

As you continue exploring the topics the participant wishes to discuss, ask targeted questions to better understand her presenting problems, life context, social support networks, self-image, and related factors. Keep in mind that you have only one session with her, and that you should make the best possible use of the information she provides in order to implement clinical strategies effectively.

*4.1.4 Preparation for the next stage of the session*

After an initial period without a set agenda, using non-directive listening, the session moves into a semi-structured phase. Mark this transition between treatment components explicitly. To do so, give a brief summary of what you have discussed so far and say that you will now guide the conversation toward specific domains of her life:

Sample summary:

*"Alright, Joana, I understand that you have been feeling quite exhausted and unmotivated lately, and that issues that previously bothered you now feel much more overwhelming. Although having a child was a dream of yours, there are times when you feel regret about motherhood. Since [BABY'S NAME] was born, it has been very difficult to balance his care with your self-care and the maintenance of essential areas of your life, such as work and social connection. Difficult moments seem to lack a solution, and in various situations, you feel hopeless…"*

Introduction to the new stage of the session:

*"To best support you right now, as I mentioned at the beginning of our session, I will need to ask you some questions about specific areas of your life. This will be very useful for the work we will do together today. From this point forward, I will introduce a few topics for us to discuss, alright?"*

You may also note that you will review the information she provided in the pre-session questionnaire if those details have not yet emerged during the initial open discussion:

*"At this point, we will revisit some of the information from the form you completed before our session, do you remember? Everything you filled out there will be helpful now, as we build upon the topics covered in those questions."*

Once you begin discussing these topics, you will be entering the second part of the session: "Assessment."

### 4.2. Part II – Assessment

This stage of the session is dedicated to gathering information that will serve as the foundation for Behavioral Activation. Here, you will explore the following topics: current routine, sources of reinforcement (current and past), social support networks, and prior experiences. The aim is to ask targeted questions about each of these domains while leaving space for the participant to add any information she considers relevant. In general, participants will already have reflected on these topics when completing the pre-session questionnaire. Because the answers on the form tend to be brief, help her expand on and explore the aspects that were not fully covered.

#### *4.2.1 Current routine*

To understand the participant's current life context, it is essential to gather information about her routine, encompassing both what she usually does and what she has stopped doing. Understanding her daily routine allows you to design individualized strategies tailored to her particular environment and to her current level of motivation to engage in meaningful activities. Begin by referring to her answers on the pre-session form, asking whether she would like to say more, and putting more specific questions about what she wrote—or about areas she may have left out.

*"I am looking at your responses from the pre-session form here. You wrote that you wake up, care for the baby, try to manage some household chores, and occasionally visit your mother, correct?*

*I see. I would like to learn a bit more about your daily routine. Could you tell me what time you usually wake up? (…) What specific household tasks do you handle?*

*You mentioned that you sometimes visit your mother. Are there other people you typically interact with day-to-day? Do you visit anyone else?"*

Below are several questions that may be useful for exploring this topic. You do not have to ask every question listed. The participant may already have covered some of these details during the initial part of the session or on the pre-session form. Focus on the aspects that have not yet been addressed or were mentioned only briefly.

**Questions about routine:**

How is your daily routine? What does a typical day look like for you?

What has changed in your routine compared to how it was before you got pregnant? (Or compared to how it was before you had the baby?)

Do you work? What are your working hours?

Do you engage in leisure activities, hobbies, etc.? Which ones?

Do you exercise?

What is the level of interaction with other people in your day-to-day life?

Does your routine include significant periods of inactivity (e.g., lying in bed, spending extended periods just scrolling on your phone, etc.)?

How is your sleep routine?

*4.2.2 Sources of reinforcement (current and past)*

In a standard behavioral activation protocol, assessment involves a comprehensive functional analysis, in which the therapist seeks to identify target behaviors, the consequences that maintain them, and the environmental contexts in which they occur (antecedents). This framework aims to map the sources of reinforcement maintaining depressive behaviors and to identify contingencies that could strengthen healthy, alternative behaviors. However, this level of assessment requires substantial time and clinical training, limiting its feasibility in brief intervention contexts. Lejuez and colleagues (2011) simplified this process without compromising its core behavioral rationale: rather than conducting a granular functional analysis, the clinician guides the participant in identifying activities aligned with her values across various life domains. In this model, values are defined as broad descriptors of how an individual wishes to live within a given domain. They serve as guiding directions that inform the selection of activities with a high probability of yielding positive reinforcement. This link to reinforcement is central: values-based activities tend to be more immediately rewarding than arbitrarily chosen tasks, thereby increasing the density of positive reinforcement in the individual's behavioral repertoire and enabling healthy behaviors to compete with and progressively displace depressive ones. The present protocol adapts this rationale into an even more streamlined structure, categorizing activities across three life domains: leisure, productivity, and relationships.

Leisure activities encompass actions that elicit feelings of pleasure, enjoyment, relaxation, or joy—that is, what the participant genuinely likes to do. Productivity, on the other hand, is associated with a sense of accomplishment; it may include tasks that do not necessarily provide immediate pleasure but foster the sense that an important goal has been met, typically yielding positive downstream consequences for the participant. Examples include studying a challenging subject, organizing the home, submitting job applications, paying bills, or resolving an interpersonal conflict. Relationships/socialization involve engaging with others and maintaining social ties. Social contact can elicit both immediate pleasure and the aforementioned sense of accomplishment. Furthermore, social interactions facilitate access to support networks that can assist during difficult periods, serving as a fundamental pillar of mental health.

Consistent with Lejuez et al. (2011), the activities identified in this protocol are not selected at random, but based on motivations that are genuinely meaningful to the participant. The core premise is that activities aligned with what the person values—because they reflect what she enjoys, considers important, or desires for her life—are significantly more likely to produce immediate feelings of pleasure, achievement, or well-being. It is precisely this reinforcing property that renders these activities therapeutically effective. By engaging in them, the participant contacts positive consequences that directly compete with the depressive cycle, facilitating the gradual restoration of a healthier behavioral repertoire. Although the term “values” is not explicitly introduced to the participant in this protocol, the underlying clinical rationale remains identical: the goal is to identify actions reflecting what matters to her, rather than assigning tasks merely to keep her busy or to fulfill external expectations.

Accordingly, aim to identify what the participant enjoys doing, what gives her a sense of productivity, and the people she values spending time with. As in the previous section, begin by referring to her answers on the pre-session form and then explore the domains that have not yet been fully addressed. Below are sample questions that can be used to explore this area:

| **Questions about sources of reinforcement** |
| --- |
| What do you enjoy doing? |
| Do you have, or have you ever had, any hobbies? |
| What do you do that helps you relax? |
| Who do you enjoy talking to? |
| What makes you feel productive? (or gives you a sense of accomplishment) |
| Do you enjoy doing activities with other people? |
| Why are these activities important to you? |
| What places do you like to visit? |

*4.2.3 Social support networks*

Help the participant identify the sources of support available to her. These include individuals, groups, and services she can turn to for help. Support networks typically involve friends, family members, co-workers, neighbors, and community groups (such as churches). You can also explore public services such as primary healthcare centers, social assistance networks, and educational institutions. Services of this kind can be particularly valuable for participants facing socioeconomic vulnerability or with limited personal support.

| **Questions about support networks:** |
| --- |
| Who can you ask for help when you need it? |
| Does any friend usually help you? |
| Does any family member usually help you? |
| Do you participate in any group (church, sports, study, etc.)? |

#### *4.2.4 Prior experiences*

Given that time is a critical resource in single-session psychotherapy, always try to build on the participant's existing resources and skills. To this end, discuss earlier periods in her life when she was not experiencing depression, or explore strategies she has used in the past to cope with challenges similar to those she currently faces. Examining these past experiences can help identify approaches that were unhelpful or made things worse, as well as those that helped her cope. Exploring this topic gives you practical ideas for her present situation—such as identifying people with whom it might be easier to reconnect, or recalling activities that were once meaningful and could be resumed. In some cases, little relevant prior experience will emerge. When that happens, the remaining protocol steps are equally valid, and you can move on to the next part of the session with the information you have gathered.

Use the following questions (Leahy, 2007)  to identify potentially useful information about the participant's history. These questions are illustrative, and it is not necessary to ask all of them:

**Questions about prior experiences:**

Before facing this current period of depression, what was your life like? What was different?

Can you try to recall a positive time in your life? What was happening during that period?

How did you cope with this issue or similar situations in the past?

Have you ever had a time when you felt isolated? How did you deal with it?

Have you ever had a time when you lacked the motivation to do things? How did you handle it?

What didn't help or got in the way when dealing with this issue in the past?

Have you ever had a time when you felt isolated? What hindered you or failed to help you deal with it?

Have you ever had a time when you lacked the motivation to do things? What hindered you or failed to help you deal with it?

| |
|---|
| Are you currently doing the things that helped you with this issue in the past? |
| Are you currently doing things that hindered you with this issue in the past? |

*4.2.5 Preparation for the next stage of the session*

Once you have gathered information on all of these topics, tell the participant that this part of the session is complete. Let her know that you will now move on to a therapeutic approach called Behavioral Activation, which has strong evidence behind it and has produced positive outcomes for many women in situations similar to hers.

### 4.3. Part III – Behavioral Activation

This portion of the session comprises: 1) explaining the rationale of the BA model, 2) delimiting the areas to be worked on and brainstorming activities, and 3) organizing the activities. The scheduling of activities will be carried out in the following stage, with the support of the Motherly app.

*4.3.1 Explaining the rationale*

Depression involves feelings of sadness, discouragement, loss of pleasure, and a decrease in activity. The model proposed by BA holds that, faced with these symptoms typical of depression, the depressed person will engage in avoidance behaviors (behaviors of isolation and avoidance of situations that may generate negative feelings) and will reduce the frequency of healthy behaviors (behaviors such as pleasurable activities, socializing activities, and productive activities, for example). The thoughts underlying this way of behaving would be something like *"I don't feel motivated, so I'm not going to meet my friend," "I don't feel like doing my work, I'll stay in bed and see whether I feel more motivated later," "I don't see the point in going out to the park like I used to, I'll stay home."* It is also very common for participants not to engage in activities and to give quite concrete justifications, describing real obstacles. In some of these cases, however, you may notice that even when solutions are proposed, participants give reasons why those solutions will not work, or describe further obstacles. All of these are cases in which an avoidance behavior is at work, and this is what characterizes the depressive cycle. The person feels low motivation and therefore stops doing activities and isolates herself; in doing so, she feels even less motivated and more discouraged, perpetuating the depressive symptoms. By isolating herself and no longer doing activities that previously produced well-being, the person with depression loses contact with the positive sensations that accompanied those moments.

Treating depression through behavioral activation seeks to break the depressive cycle. Even if the person feels sadness, discouragement, and lack of will, she should gradually engage in activities that may generate sensations of well-being, self-efficacy, pleasure, and so forth. Years of research on BA show that: **changing the way you act can change the way you feel.** Although engaging in activities may be experienced as difficult, tiring, or demotivating at first, the therapeutic effect of BA comes precisely from this effort to change behavior in order to then feel the change in the sensations we experience. In short, help the participant identify actions she has stopped doing or is avoiding because of depression, and take the first steps toward resuming them, building a more active and healthy routine.

Below is a brief model of how the rationale may be explained:

*"Many people go through periods in which they feel sadder, less motivated, more pessimistic. This can happen without an apparent reason or after a very difficult event. Sometimes there may even be difficulty feeling pleasure in things that were previously considered enjoyable."*

*"The tendency during these periods is to avoid situations, places, or people that could bring up bad feelings. In some cases, people even stop doing many things they used to do because this avoids bad feelings. In the short term, this strategy may bring relief, but it can make the situation worse after a while. By avoiding situations because they bring bad feelings, you lose contact with important people (friends, family), with resources, opportunities, and other essential things. This creates more frustration, lack of motivation, sadness, and other negative feelings. In other words, a vicious cycle is created."*

*"From your answers in the form and from what we have talked about so far, it seems that you are going through something similar. Our goal is to help break the vicious cycle that keeps you feeling this way. The proposal is to help you engage in activities that are important to you and that can make you feel good. The approach we are going to use is called* ***behavioral activation****. According to this approach, well-being is associated with a routine of healthy behaviors, that is, behaviors that allow you* ***more quality of life and bring you closer to what is important to you****."*

*"It is very natural to think that our actions are guided by our feelings. For example, I feel unmotivated and therefore I stop doing certain things. In that case, it is even more natural to think that in order to go back to doing the things I stopped doing, I need to change my*

*motivation first. But scientific research shows that the opposite path also works: if we change the way we behave, we can change our feelings and thoughts. If we regularly engage in activities that are important to us, we can change how we feel and think. It is harder to feel sad or down if our actions bring a feeling of pleasure and accomplishment."*

##### *4.3.2 Delimiting the areas to be worked on and brainstorming activities*

After explaining the BA rationale and addressing any questions the participant may have, help her define the areas that will be worked on and the possible activities she can do or resume as part of therapy. Keep in mind three important areas to cover with the participant: leisure, productivity, and relationships.

*"Let's try to think of activities that can make you feel better. The idea is to identify and plan daily activities within three areas:* ***leisure****, which includes activities you do to relax or have fun;* ***productivity****, which includes obligations in general and habits that may be healthy for you; and* ***relationships****, which is spending time with the people around you."*

The information you generate here will be used by the participant to schedule activities over the coming weeks. It is recommended that you make a list (in Google Docs, a notepad app, or another tool) and share your screen so that the ideas can be written down as they come up. If the participant has difficulty thinking of activities, one strategy that can help is to ask what she used to enjoy doing at other points in her life and is no longer doing. You can ask her, for example, about activities she used to do and stopped, people she likes but has not kept in close contact with, and actions that made her feel productive (or revisit these, if they have already been discussed).

Activities should not be thought of merely as ways of keeping the participant busy, but as actions that matter within what she, as an individual, believes to be important. Seek to understand whether that activity is in line with how the participant would like to live or with what she values. You can ask questions such as:

*"Do you feel this activity is important to you, not because other people think so, but because you yourself consider it important?", "What kinds of activities do you feel, in your heart, that you would like to do?", "Try to think about what you would like to practice in your life because it is important to you. What comes to mind?", "What is depression preventing you from doing that is important to you?"*

If you perceive that the tasks suggested by the participant are exclusively ways of meeting other people's expectations, you can ask:

*“How motivated are you to do this activity?”, “Why is this task important to you?”, “Do you think that if no one were asking it of you, it would still be important?”, “If no one were asking anything of you, what kinds of things would you like to do more of in your daily life?”*

By the end of this stage, the idea is to have a list of activities that are in some way important, enjoyable, or valuable to the participant.

*4.3.3 Organizing the activities*

With the list of activities in hand, begin organizing the ideas you came up with. This phase will be important for promoting the participant’s engagement. The participant is not expected to carry out every activity on the list. Select activities following the criteria explained below. The chances of the participant putting into practice what you agree upon will be significantly higher if the activities are planned according to these criteria.

1) **Rank them by level of difficulty:** the listed activities should be classified as easy, medium, or hard. It is important to select activities of varying degrees of difficulty. Instruct the participant that as she completes activities of lower difficulty, she may gradually increase the difficulty level of her planning. For example:

*“When planning an activity, it is important to think about how difficult it may be. There are three **levels of difficulty**: **easy**, which are those activities you think you could do quickly and without effort; **medium**, which are those that require more effort; and **hard**, which are the more challenging ones. To increase the chances of success, it is important to select activities of varying degrees of difficulty and to start with the easiest ones. Shall we classify these activities from the list according to their level of difficulty? I see here that we wrote down (…). How difficult do you think it will be to carry out [activity]? As you meet your goals, you can start trying the harder ones.”*

2) **Being feasible:** the increase in activity level should be substantial yet gradual. To this end, take into account how active the participant’s current routine is and set goals that are achievable in her context. Larger goals should be broken down into smaller but attainable targets. For instance, a full house cleaning can be broken down into smaller activities such as sorting the laundry, tidying one room, organizing one cupboard, or sweeping the living room. Each of these smaller tasks could be placed on a different day, or in a different part of the day. For example:

*“Something that can help is trying to ‘break’ activities into small parts. For example, if you want to organize your bedroom, you can start with small sections (a drawer or a cupboard). Once you manage to reach that small goal, you move on to the next one. The simpler the activity, the easier it is to succeed at it. Is there any activity you think it would make sense for us to break into smaller parts? (…) That activity seems challenging—how about we divide it into small steps?”*

3) **Identifying people who can help, if any:** given the motivation-related difficulties that come with depression, asking other people for support can help. Identifying people who can join her in an activity, or assist her with it, is a way of increasing the chance of success and of creating moments of social contact at the same time. For example:

*“The chances of overcoming feelings of sadness, lack of pleasure, or lack of motivation increase if you have the support of other people. Involving other people in your activities can make everything more enjoyable. At the same time, it can improve your relationship with that person. All of this can have a positive impact on your well-being.”*

*“From among the people we identified earlier, try to choose someone you trust whom you could invite or ask for help. For example, if you find grocery shopping very tedious, someone’s company can make the experience more pleasant. One tip is to think about how that person could help you. You can recall the people we thought of earlier in the session. Who do you think could help you with the activity or activities we listed here? Let’s write that person’s name next to the activity together.”*

*“When you ask for help, you can explain to the person exactly what they could do to help you. Just remember that the idea is for this person to enable you to do something, not to do it for you.”*

By the end of this stage, you and the participant will have a list of activities organized by level of difficulty, with support people identified where relevant. With this list in hand, move on to Part IV, in which the Motherly app will be introduced and the activities will be scheduled.

### <u>4.4. Part IV – Motherly app</u>

After randomization, the participant will receive a video explaining how to install Motherly and log in. She is therefore expected to already have the app ready to use on her phone. It is important, however, to be prepared for the case in which this has not happened. If so, help the participant until she is able to open the app, and then go on to schedule the behavioral activation activities.

This is the link to the video that will be made available to the participant beforehand. It is essential that you watch it, so that you can assist her with the app installation process in case she has not watched the video previously or has questions at any stage of the download (https://youtu.be/SMfUfHxOq58).

*"Alright, let's do that now, then. The app is called Motherly; you can search for it in the Play Store (…) Now that you have it installed, you will need to create an account. (…) When you get to the home screen, you can tap the"Create activity" option. A screen will open for you to write which activity you are scheduling, which area of life it belongs to, its difficulty level, and the date and time to do that activity."*

As soon as the participant has the app open, transition to scheduling:

*"Now that you have the app open, let's use it to schedule the activities we planned together. After we do that, I'll show you the other features it has."*

#### *4.4.1 Scheduling the behavioral activation activities*

The study management team recorded a second video showing, in practice, how to create a behavioral activation activity in the app. After the psychotherapy session, you will thank the participant for the time you spent together and send her this material, so that she can watch it and refer back to it whenever a question comes up while using the app (https://youtu.be/7pfAWjUx8wM?si=C_Idz1kFVY5VfkvS).

Drawing on the content discussed in the session so far and on the list prepared, help the participant use the Motherly interface to schedule activities. It is recommended that activities be scheduled for the next 2 weeks. As a general rule, choose together with the participant one activity at each difficulty level (easy, medium, and hard). Remember that for activities considered hard it may be useful to break them into smaller parts.

If the participant shows resistance to certain activities, there are a few things you can do. If the reason is the perceived difficulty of the activity, consider breaking it into even smaller parts. You can also revisit the people or support networks she can ask for help. While scheduling the activities, it may be important to return to the BA rationale: show that you understand her lack of motivation, but remind her that one of the ways to feel better is by doing activities that are pleasurable and/or important to her. In any case, if the participant says she does not feel ready to carry out an activity, prioritize others; she can schedule that one herself later on.

*"I see, that activity seems too difficult. Perhaps it did not seem so difficult when we were putting the list together. That can indeed happen; it may be that the barriers to doing it have become clearer now. If you do not feel* ***ready*** *to do this right now, you can wait a few weeks to start. Another thing we can do is divide this activity into smaller parts that seem more doable. What do you think?*

*(…)*

*It is always important to remember that by doing the activities you can feel better. Remember what I talked to you about earlier? In depression it is normal to feel less willing to do a number of things. But by resuming a routine with activities that make you feel good and* ***generate*** *well-being, feelings gradually change and little by little you can feel better. How about trying to do that?"*

Explain to the participant that she should continue scheduling and doing activities over the next four weeks. Briefly revisit the rationale, explaining the effect of a more active routine on the sensations associated with depression. Make sure she understands how activity scheduling works in the app and address any questions she may have.

*"Great! We have now finished scheduling your first behavioral activation steps. We started this process together in the session, and over the coming weeks you will carry it forward by doing something similar to what we did, with the help of the app. You just need to remember what we did here. I will send you this list of activities we already have; you can update it with others if you have new ideas, or gradually implement the things we thought of. It is natural that not every activity will turn out as we planned, but we don't need perfectionism here. Try to put into practice the things we talked about, and keep scheduling activities and putting them into practice over the next four weeks."*

##### *4.4.2 Explaining the Motherly app's other features*

Before closing the part of the session dedicated to Motherly, show the app's other features. Place greater emphasis on explaining those modules you judge to be related to the participant's needs (e.g., if the participant spoke about insomnia, show the module on sleep).

4.5. Part V – Closing

After the interventions proposed in this protocol have been carried out, you will close the single-session psychotherapy. Communicate to the participant that the session is coming to an end. Provide a brief summary of the main points addressed, such as the problems discussed, the logic of behavioral activation, the support networks, and the next steps regarding the Motherly app. Emphasize the importance of continuing to use Motherly over the coming weeks as part of the treatment.

Emphasizing the importance of Motherly and opening space for questions:

*"You now have a part of the treatment that will take place over the coming weeks. The idea is for you to keep putting into practice what we discussed during the session. As you saw, the app will help you with that. You already understand how it works; just keep scheduling activities and putting them into practice, and you will be taking important steps toward caring for yourself at this time. Do you have any questions about what we talked about? Is there any point that was not very clear or that you would like me to explain better?"*

Before ending the video call, tell the participant that she still has three stages ahead of her in the study. Explain that in the next stage the study management team will send her questionnaires via WhatsApp for her to answer online. After that, someone will contact her again to schedule the final assessment with the assessor. Finally, one last questionnaire will be sent for her to answer online.

Let the participant know that these final study stages will take approximately one month in total. Reassure her that she need not worry, as all instructions and communications will come via WhatsApp throughout this period, and that the team will remain available to answer any questions.

Conversely, in cases where the safety plan was worked on instead of behavioral activation and the other components, the closing is different. Provide the summary by revisiting the points listed in the safety plan and the importance of attending the appointment with the professional to whom she is referred.

*"We talked about ways to keep you safe in the face of this difficult moment and of the thoughts you have been experiencing. Keep in mind what we discussed, and if you go through a crisis, open this plan and follow it step by step, okay? As we agreed, once we finish here you are going to print it and leave it on the table (or whichever other option you* ***have*** *agreed on), right?* ***In addition, someone from our team will contact you to assist you.*** *It will be very important that you attend the appointment with her. Do you have any*

*questions about what we talked about? Is there any point that was not very clear or that you would like me to explain better?"*

### 5. Guidance for psychiatric emergencies and domestic violence

If you observe that the participant is at risk—whether due to suicidality, urgent situations, or violence—make a referral to specialized services that can provide the appropriate level of care. Specific guidance on higher-risk cases is provided below.

#### *5.1. Psychiatric emergencies: suicidality*

One of the study's exclusion criteria is the presence of suicidal ideation. Participants were therefore assessed for this symptom during the recruitment/assessment process, and there is no need to conduct a suicidality assessment with every participant. Nevertheless, it is possible that some of them may present signs of suicidal ideation and/or behavior (statements that she no longer feels like living, that things would be easier if she were not alive, that she would rather not have been born, etc.). In such cases, address the subject directly.

Provide a welcoming, non-judgmental context; do not minimize or invalidate the problem, and do not display shock. Remain calm and ask, through direct questions, whether the participant is thinking about suicide: *"Are you experiencing suicidal thoughts?", "Are you thinking about taking your own life?"*

If the participant confirms that she is currently experiencing suicidal ideation or planning, **you must not proceed with the components of this protocol, such as behavioral activation and use of the app. The focus shifts to the suicidal behavior.** You must: 1) build a safety plan with her, and 2) provide a referral to specialized services.

Explain to the participant what the two of you will do. Tell her that, because of the thoughts she is facing, you will build a plan that can help her deal with the problem. This planning involves a safety plan and a referral for a psychiatric consultation.

The safety plan is a technique aimed at helping participants recognize when a crisis is occurring and what can be done during crises so that they remain safe. This involves listing things she can do to distract herself, people she can ask for help, professionals she can contact, and other strategies. Write the plan down. Open a document (Google Docs, Word, etc.), share your screen, and write the safety plan according to what is discussed with the participant. When you finish writing it, send this document to her and discuss how to ensure it is easily accessible if she needs it (e.g., printing it and leaving it somewhere, keeping a screenshot on WhatsApp, copying it into a notes app, etc.).

| Safety Plan Structure | |
|---|---|
| What triggers the desire to kill myself or to harm myself? | |
| How can I recognize that I need to take the first steps to look after myself and keep myself safe? | |
| When I notice that one of these things has happened, or when I feel that I want to die or to hurt myself, I will do the following: | |
| Remove the means I have already used, or might come to use, to hurt myself. I will also ask someone I trust to help me with this: | |
| I will try to relax by: | |
| I will do some physical activity, for example: | |
| I will divert my attention by: | |
| I will repeat the following thoughts to myself: | |
| I will get in touch with people who give me strength: | Name, phone |
| I will call my psychiatrist and my psychotherapist: | Name, phone |
| Things that make it worth staying alive: | |
| Date and signatures:<br><br>participant and professional | |

Once the safety plan is complete, make the referral. In this study, we will have psychiatrists who will receive cases involving suicide risk. You will be given the contact details of the professional to whom you should make the referral. Explain the reason for the referral to the participant and emphasize the importance of her contacting this professional.

**Notice:** It is of fundamental importance that cases of suicide risk be reported to the clinical supervisors (Dr. Pedro Zuccolo, [phone]; or Dr. Daniel Fatori, [phone]).

*5.2. Cases of domestic and family violence against women*

If signs or reports of domestic violence arise during the session, it is essential to approach the matter with sensitivity, respect, and a focus on the participant's safety. Whenever possible, ask permission before exploring the topic in greater depth, for example: *"May I ask you some questions about this so that I can better understand how you are doing and think about how I can help you?"*

Prioritize active listening and an empathetic, non-judgmental stance, recognizing that talking about violence can be extremely difficult and ambivalent. It may be useful to communicate this understanding explicitly, for example: *"I imagine it may be very difficult to talk about this. Thank you for sharing it with me."*

It is also important to validate the participant's experience, reinforcing that violence is unacceptable and that she is not responsible for the aggressive behavior directed at her. Phrases such as *"What you are describing is a form of violence, and you do not deserve to be treated this way"* and *"This is not your fault"* can help reduce the feelings of guilt and shame that are very common in these situations.

It is also important to recognize that many women remain in violent relationships for multiple reasons, such as fear, economic dependence, concern for their children, a desire to preserve the relationship, lack of social support, or difficulty identifying safe alternatives, among others. These factors should be understood with empathy, avoiding any kind of judgment.

During the session, name the situation as violence, validate the emotional ambivalence experienced by the participant, assess the available support network (such as family members and close friends), and explore possible courses of action without pressuring the participant to make a decision—for example, identifying a safe place and listing emergency contacts and support services. Since ending a relationship does not automatically guarantee an end to the violence, and may in fact represent a moment of heightened risk, it is important that this process be carried out with caution and planning.

In this regard, plan collaboratively with the participant which women's service networks or family and social support networks she has already used at some point in her life or is already familiar with in her region (see the list below). If the participant does not identify these service networks, it may be useful to check whether there are services available in her region that can be accessed safely and feasibly. You may take a more active role by presenting the support networks (listed below) to the participant, always checking whether the suggestion makes sense for her reality. Guide this topic with the following questions:

*“In difficult moments in the past, what helped you deal with the situation?”*

*“Is there anyone, or any place, where you feel safe talking and asking for help?”*

*“In your city, is there any place that assists women going through this situation, such as women’s police stations, referral centers, or health units?”*

It is worth emphasizing that situations of violence must be properly documented and taken to clinical supervision, given the complexity and the ethical issues involved in managing these cases.

**Support network for women:**

***In Public Security:***

- General and specialized police stations (*Delegacias Especializadas de Atendimento à Mulher*, DEAMs — police stations specialized in offences against women);
- Regular Military Police patrols (*Polícia Militar*) and *Patrulha Maria da Penha* patrols (units dedicated to monitoring women under protective court orders);
- Regular Municipal Guards and *Guarda Maria da Penha* units;
- Fire Department (*Corpo de Bombeiros*);
- Forensic Medical Institute (*Instituto Médico Legal*, IML).

***In the Justice System:***

- General and specialized courts;
- General and specialized prosecutor’s offices (*Promotorias*);
- Public defender’s offices (*Defensorias Públicas*) and their Women’s Defense Units (*Núcleos de Defesa da Mulher*).

***In Health:***

- Health posts and health centers (*Postos/Centros de Saúde*);
- Hospitals;
- Health services specialized in caring for women in situations of violence;

- Mobile emergency care service (*SAMU – Serviço de Atendimento Móvel de Urgência*).

***In Social Assistance:***

- Shelters (*Casas-Abrigo*);
- Reception houses for women in situations of violence (*Casas de Acolhimento para Mulheres em Situação de Violência*);
- Specialized Social Assistance Referral Centers (*Centros de Referência Especializados de Assistência Social*, CREAS);
- Social Assistance Referral Centers (*Centros de Referência de Assistência Social*, CRAS).

***In addition, women can also count on:***

- *Casas da Mulher Brasileira* (one-stop centers integrating several women's services under one roof);
- Women's referral and specialized care centers (*Centros de Referência/Especializados de Atendimento à Mulher*);
- Mobile units for women's assistance (*Unidades Móveis de Atendimento à Mulher*);
- *Ligue 180* — the national toll-free helpline for women.

### 6. Guidance on common complaints, additional techniques, and management

Although each woman is unique in her experience, previous studies conducted by our research group have revealed recurring themes among women with postpartum depression. The following section describes some of the clinical complaints most commonly identified in those studies, together with guidance on how to address them within the context of the proposed intervention. The aim is to offer suggestions that facilitate clinical management when these issues are identified. This section is therefore not a fixed script to be followed, but a flexible guide that can be adapted according to the needs and particularities of each participant.

#### *6.1. Social support issues*

Social support plays a central role in coping with depressive symptoms in the postpartum period. Women with PPD benefit from the support of partners, family members, and broader social networks—such as friends, neighbors, co-workers, and the community—which can help them adapt to motherhood and deal with emotional distress.

Social isolation, in turn, is a risk factor that contributes to the worsening of maternal mental health. Even when there are people close by, many women may still report feelings of loneliness, frequently associated with fear of judgment, the stigmatization of depression, or the perception of having no available bonds.

The absence or fragility of support, such as absent partners or difficulty asking for help, can further intensify depressive symptoms and increase the burden of infant care. It is worth noting that this isolation may arise both from the absence of intimate relationships and from a lack of integration into a broader social network. Although it is a complaint common to most mothers, socially marginalized groups tend to be more vulnerable to this phenomenon.

If this is identified as an issue, list with the participant the support network currently available to her. To this end, it may be useful to ask what her relationships were like before the depression and who her closest people were. If such people exist, seek to identify them and discuss the possibility of behavioral activation focused on social engagement—for example: arranging to go out with a friend, calling her parents, or resuming close personal conversations.

If the participant perceives that her support network is limited or nonexistent, validate the emotional experience associated with this and acknowledge that building bonds can be a gradual process. In such cases, the focus of behavioral activation may initially be on increasing exposure to social contexts, even without the immediate expectation of establishing close relationships. Seek to help her identify opportunities to build or resume bonds, which can also be a behavioral activation activity—for example: visiting public or collective spaces (squares, libraries, street markets, groups), taking part in structured activities (mothers' groups, short courses, activities at health units, gatherings at religious or community centers), or interacting in moderated online spaces, such as mothers' communities.

#### *6.2. Low communication skills*

Difficulties with communication skills are frequent among women with PPD and can compromise both emotional expression and the ability to ask for help. Many mothers report difficulty verbalizing feelings related to motherhood, such as ambivalence, frustration, or exhaustion, as well as difficulty making clear requests regarding their needs to partners, family members, or their support network. This pattern can contribute to increased emotional distress, reinforcing feelings of being misunderstood, lonely, and overburdened.

If this need is identified, help the participant identify and name the specific needs she would like to communicate—for example: the need for help with household tasks, with infant care, for company, and for encouragement. On that basis, it is important to validate these needs, highlighting that asking for help and expressing feelings is a legitimate behavior in the face of demanding emotional circumstances.

You can briefly explain the three main types of communication: passive, aggressive, and assertive. Passive communication occurs when a person has difficulty expressing her own needs, feelings, or opinions, frequently prioritizing the needs of others and avoiding conflict. Aggressive communication, in turn, involves expressing one's own needs in an imposing or hostile way, potentially disregarding or disrespecting the feelings and rights of others. Assertive communication, in contrast, consists of expressing thoughts, feelings, and needs in a clear, direct, and respectful manner, taking into account both one's own rights and those of the other person. Ask the participant how she imagines each of these ways of communicating the previously identified need would look. You can help her develop examples of assertive communication and reinforce the importance of assertive communication as a healthy communication style for relationships.

A brief behavioral rehearsal may also be proposed, in which you play the role of the participant, modeling how to communicate needs clearly and assertively. The roles can then be reversed so that the participant practices this form of communication, and you can provide corrective and reinforcing feedback when necessary.

Finally, it may be useful to plan this communication as a behavioral activation activity, helping to define concretely when, where, and with whom she can have this conversation—for example: asking a family member to look after the baby for a period while she is out of the house. The important thing is for the participant to understand that her needs are valid, that it is possible to communicate and stand up for her needs without attacking the other person, and that planning these interactions concretely can encourage her not to avoid emotionally difficult conversations.

#### *6.3. Identity issues*

Many women with PPD feel absorbed by the role of mother, as if this had become the only aspect of their identity. As a result, activities, interests, and roles that were previously sources of pleasure and meaning may end up being set aside. The consequences of this may appear as intense and frequent feelings of guilt, failure, or inadequacy.

If this need is identified, explore with the participant aspects of her identity before motherhood, investigating which activities, interests, and topics were part of her routine and which were emotionally meaningful to her. It is useful to understand how the participant felt when carrying out these activities and what role they play in her sense of identity and well-being.

Regarding the emotions associated with this issue, such as guilt, frustration, or a sense of having lost herself, validate that motherhood involves important changes in routine, in priorities, and in the way time is organized. It is therefore common for some previously valued activities to take up less space in daily life. At the same time, you can reinforce that maintaining some contact with these parts of her identity can be important for her well-being, even if in an adapted form.

In this sense, it may be useful to encourage the participant to gradually resume activities that were meaningful to her, adjusting expectations regarding frequency, duration, or performance. Small resumptions—such as devoting a short period of the week to a personal interest, or adapting a former activity to her current reality—can be planned as behavioral activation activities, prioritizing concrete, feasible actions that are compatible with the participant’s current stage of life.

*6.4. Maternal self-efficacy issues*

Issues related to maternal self-efficacy are frequent in women with PPD, especially during the initial period of adaptation to motherhood or with babies of difficult temperament. In these situations, many mothers may doubt their own ability to care for the baby, interpreting difficulties as personal failures. The burden may increase as a result of sleep deprivation and constant demands, further intensifying feelings of exhaustion, irritability, and hopelessness, and reinforcing beliefs that they are bad mothers.

If this thought pattern is identified, use Socratic questioning to help the participant make very rigid or extreme interpretations of her maternal competence more flexible. Initially, it is important to identify the most relevant thought associated with the emotional distress, asking the participant what usually goes through her mind when she feels bad about herself as a mother. From there, the aim is to identify the central thought that most influences this emotional state—for example: *"I am a bad mother"* or *"I don't know how to take care of my baby."*

Next, explore with the participant what evidence supports this thought and what contradicts it, and reflect on the impact this way of thinking has on her well-being and on the extent to which it actually helps her deal with the situation. Explore, too, the advantages and disadvantages of holding on to this kind of interpretation.

Based on this analysis, you and the participant can jointly construct a more balanced and realistic alternative interpretation of the situation. One strategy that can help in this process is to invite the participant to imagine that someone she loves were facing the same difficulty and to reflect on what she would say to that person. This exercise tends to favor more compassionate and less self-critical responses. For example: *"I am having difficulty getting my child to fall asleep on his own, but that does not mean I am a bad mother. This is a common difficulty among many mothers. Right now I am facing challenges, but I can learn new strategies and ask for help when necessary."*

At this point, guide the participant to use the section of the app under "Health" that relates to her main difficulty. Sleep and stress management issues, for example, can be addressed through the "Sleep hygiene" module under "Relaxation Sessions" and "Coping with stress" under the "Mental Health" section. Indeed, accessing and completing these modules can be an activity scheduled for behavioral activation. To this end, it is important that you show the participant where to find the section in the app and that you create the activity collaboratively in the app.

*6.5. Infant care*

Problems related to the baby's sleep, feeding, and breastfeeding are frequent complaints and can generate exhaustion, insecurity, and feelings of maternal incapacity. Sleep deprivation and uncertainty about caring for the baby "the right way" tend to increase anxiety and emotional burden, especially when these difficulties persist or when the mother perceives herself as lacking adequate support. These experiences, although common, are usually gone through in isolation and can contribute to the worsening of psychological distress, especially in women with depressive symptoms in the postpartum period.

If these needs arise, validate the impact of these difficulties, acknowledging how draining issues related to sleep, feeding, and infant care can be, both physically and emotionally. It may also be useful to normalize the fact that many of these challenges are common in the postpartum period and are part of the process of adapting to a routine with the baby.

In addition, guide the participant to access the psychoeducational content available in the library, in the app's postnatal section, which addresses topics such as infant sleep, feeding, breastfeeding, and practical strategies for dealing with these situations—for example, the modules "Mother's life," "Baby development," and "Breastfeeding." Access to evidence-based information can help reduce uncertainty, broaden the repertoire of strategies, and strengthen the sense of maternal competence. When relevant, briefly discuss this content during the session, helping the participant reflect on which guidance may be most useful and applicable to her reality.

*6.6. Career crisis triggered by motherhood*

Motherhood can give rise to significant professional dilemmas, especially when the mother experiences insecurity or a loss of motivation to resume her career. Cases of the "triple shift" are also common, in which the mother juggles paid work, infant care, and household responsibilities, which can generate exhaustion, reduced performance, and emotional distress.

Many mothers may report difficulty recognizing themselves professionally after the birth of the baby, feeling disconnected from their previous life, their competencies, and their former relationships, especially during maternity leave or in cases in which they temporarily interrupt work.

In practice, you can help the participant build a gradual and realistic plan for returning to work, exploring concrete aspects of this transition. This may include reflecting on when a feasible moment to return to work would be, what the first steps would be, and who could help with infant care during this period—such as a partner, family members, a daycare center, or private caregivers. To this end, you can use the structure of the activity organization and planning strategies described in Part III of Chapter 3 on Behavioral Activation.

Validate the ambivalent emotions that may arise in this process, such as guilt, fear, and insecurity, alongside the desire to resume parts of her professional life, reinforcing that these feelings are common in the transition to motherhood.

If work is something the participant values, highlight the importance of preserving this aspect of her identity, recognizing that professional life can also contribute to a sense of personal accomplishment, autonomy, well-being, and care for her children. When there is doubt or low motivation about returning to work, explore the advantages and disadvantages of resuming or starting professional life at this moment, helping the participant reflect more clearly on what makes the most sense for her stage of life and which paths may be more consistent with her values and needs.
